# Identification of Digital Twins to Guide Interpretable AI for Diagnosis and Prognosis in Heart Failure

**DOI:** 10.1101/2024.11.11.24317106

**Authors:** Feng Gu, Andrew J. Meyer, Filip Ježek, Shuangdi Zhang, Tonimarie Catalan, Alexandria Miller, Noah Schenk, Victoria Sturgess, Domingo Uceda, Rui Li, Emily Wittrup, Xinwei Hua, Brian E. Carlson, Yi-Da Tang, Farhan Raza, Kayvan Najarian, Scott L. Hummel, Daniel A. Beard

## Abstract

**Background:** Heart failure (HF) is a highly heterogeneous and complex condition. Although patient care generates vast amounts of clinical data, robust methods to synthesize available data for individualized management are lacking.

**Methods:** A mechanistic computational model of cardiac and cardiovascular system mechanics was identified for each individual in a cohort of 343 patients with HF. The identified *digital twins*— comprising optimized sets of parameters and corresponding simulations of cardiovascular system function—for patients with HF in the cohort is used to inform both supervised and unsupervised approaches in identifying phenogroups and novel mechanistic drivers of cardiovascular death risk.

**Findings:** The integration of digital twins into AI-based analyses of patient data enhances the performance and interpretability of prognostics AI models. Prognostics AI models trained with digital twin features are more generalizable than models trained with only clinical variables, as evaluated using an independent prospective cohort. In addition, the digital twin-based approach to phenomapping and predictive AI helps address inconsistencies and inaccuracies in clinical measurements, enables imputation of missing data, and estimates functional parameters that are otherwise unmeasurable directly. This approach provides a more comprehensive and accurate representation of the patient’s disease state than raw clinical data alone.

**Interpretation:** The developed and validated digital twin-based AI framework has the potential to simulate patient-specific pathophysiologic parameters, thereby informing prognosis and guiding therapeutic options.Ultimately, this approach has the potential to enhance the ability to focus on the most critical aspects of a patient’s condition, leading to individualized care and management.

**Funding:** National Institutes of Health and Joint Institute for Translational and Clinical Research (University of Michigan and Peking University Health Science Center)

## Introduction

Heart failure (HF), both with preserved and with reduced ejection fraction (EF), is a heterogeneous syndrome characterized by abnormal cardiac structure and function, leading to reduced cardiac output and/or elevated filling pressures at rest or with exertion [1, 2]. This highly prevalent multifactorial condition remains associated with significant morbidity and mortality [3]. A major challenge in HF management is individualization of risk and treatment, with relatively fewer adequate treatment options for those withheart failure with preserved ejection fraction (HFpEF) than for those with heart failure with reduced ejection fraction (HFrEF) [4–6]. Due to the wide range of clinical presentations and diverse underlying mechanisms, it is often difficult to tailor treatments to individual needs [7].

To better understand and assess heterogeneity in HF, unsupervised machine learning (ML) has been applied to patient record data to identify patient subgroups with similar characteristics that have distinct prognoses [8–12]. Recent applications integrate molecular biomarkers with clinical variables to enhance mechanistic insights into phenogroups identified through ML [13–15]. Nonetheless, although unsupervised ML-driven analyses of large HF patient datasets have successfully identified phenogroups with different prognoses, interpreting the mechanisms behind these differences remains limited and relies on prior knowledge. Similarly, supervised learning approaches, either using traditional ML or deep learning, can be trained to robustly predict mortality in HF patients [16–18]. However, supervised ML models used to improve performance do not easily yield mechanistic interpretation.

We have developed a model-based, physiology-informed ML classification approach to categorize HF patients into distinct phenotypic groups, revealing that when explicit physiological prior knowledge embedded in a computational model is integrated with data-driven inference, the phenotyping process is enriched compared to a purely datadriven approach [19]. Here we build on that framework utilizing digital twins—comprising optimized sets of parameters and corresponding simulations of cardiovascular system function—for HF patients in a retrospective cohort to inform both supervised and unsupervised approaches in identifying phenogroups and novel mechanistic drivers of cardiovascular death risk. We demonstrate how the integration of digital twins into AI-based analyses of patient data enhances the performance and interpretability of prognostics AI models. Finally, we show that prognostics AI models trained with digital twin features are more generalizable than models trained with only clinical variables, as evaluated using an independent prospective cohort.

## Methods

### Study design and patient data extraction

A retrospective cohort of 343 patients, selected from all patients with HF who visited the University of Michigan Health System (UMHS) between June 1, 2009, and November 30, 2023, was identified using Electronic Health Records (EHR) data. Cohort discovery is briefly summarized in Figure 1. Another independent prospective cohort of 86 patients with dyspnea from the University of Wisconsin (UW)-Madison was also included for validation of the prognostic model. This cohort includes 9, 49, 6, and 5 patients in pulmonary hypertension (PH) groups 1, 2, 3, and 4, respectively, and 17 patients that presented with idiopathic dyspnea. Of those in group 2 PH, 32 were diagnosed with HFpEF. Further details are provided in the supplementary material *(Method S1)*.

**Figure 1:**
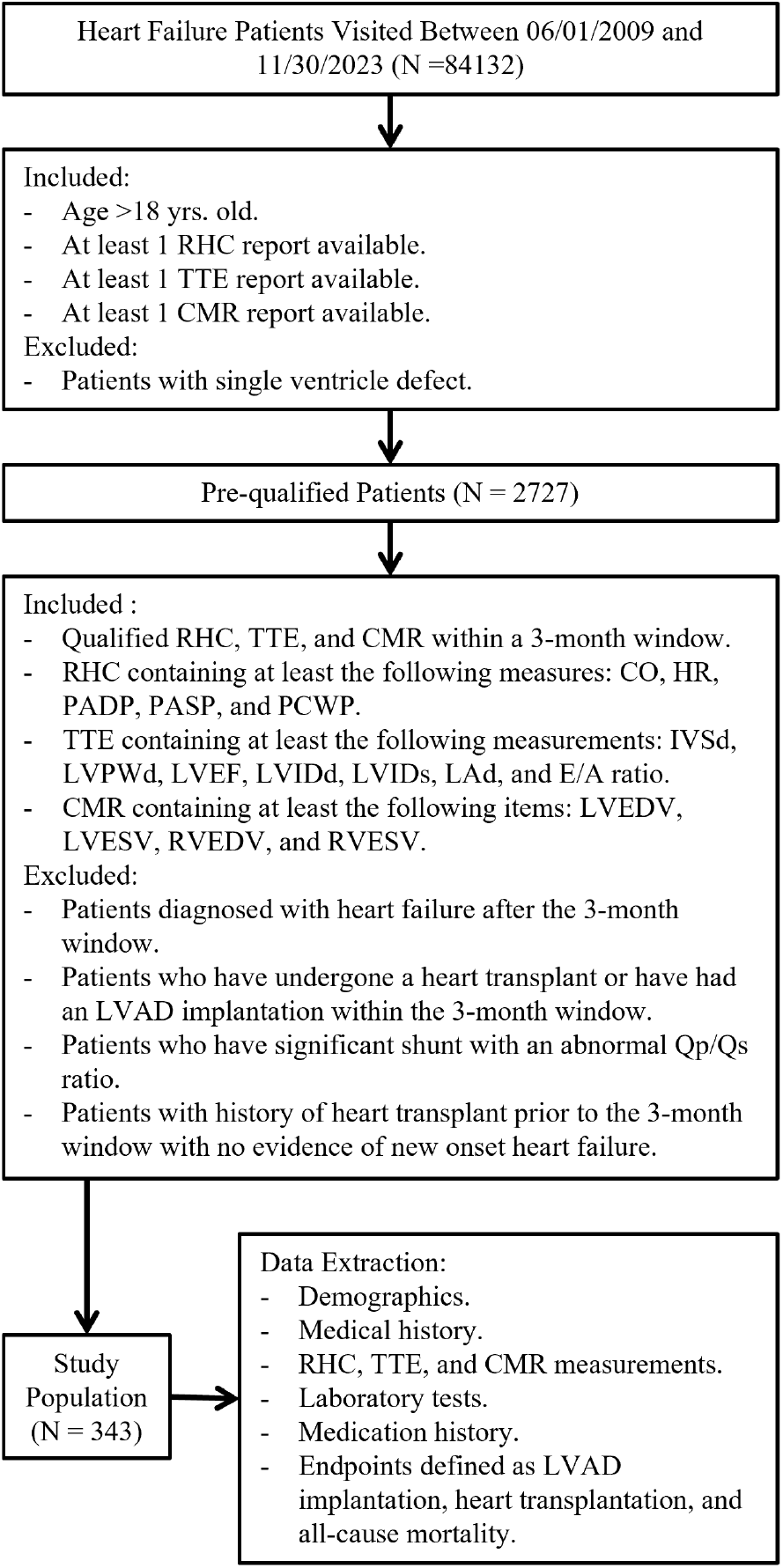
UMHS cohort discovery. The flowchart represents the University of Michigan Healthcare System cohort discovery process. The extracted data are used for digital twin identification, precision phenotyping, and building prognostic AI models. RHC = Right Heart Catheterization, TTE = Transthoracic Echocardiography, CMR = Cardiac Magnetic Resonance Imaging, CO = Cardiac Output, HR = Heart Rate, PADP = Pulmonary Artery Diastolic Pressure, PASP = Pulmonary Artery Systolic Pressure, PCWP = Pulmonary Capillary Wedge Pressure, IVSd = Interventricular Septum Thickness, LVPWd = Left Ventricular Posterior Wall Thickness, LVEF = Left Ventricular Ejection Fraction, LVIDd = Left Ventricular Internal Diameter end-Diastole, LVIDs = Left Ventricular Internal Diameter end-Systole, LAd = Left Atrial Diameter, LVEDV = Left Ventricular End-Diastolic Volume, LVESV = Left Ventricular End-Systolic Volume, RVEDV = Right Ventricular EndDiastolic Volume, RVESV = Right Ventricular End-Systolic Volume, Qp/Qs ratio = the ratio of pulmonary blood flow (Qp) to systemic blood flow (Qs), LVAD = Left Ventricular Assist Device.

### Identification of digital twins

We created parameterized representations of each patient using a computational model of the cardiovascular system by matching simulation outputs to corresponding EHR data. The individualized model for a given patient— comprising an optimized set of model parameters and corresponding cardiovascular simulations—constitutes the patient’s digital twin. The computational model used for simulating digital twins includes a central and peripheral parts (Figure 2A). The central part, representing the heart, is based on the three-wall segment (TriSeg) heart model of Lumens *et al*. [20](Figure 2B). The TriSeg model is coupled to lumped elements representing the peripheral circulation, atria, and pericardium. This model represents an optimal balance between maintaining relatively low computational complexity and achieving sufficient detail to reflect key anatomical features assessed by TTE and CMR. The peripheral part of the model is coupled to the TriSeg heart model, forming a closed-loop model and enabling the simulation of blood flow and pressure dynamics throughout the cardiovascular system (Figure 2C). A detailed description of digital twins identification is provided in the supplementary material *(Method S2)*. The MATLAB (MathWorks Inc.) source code for the algorithm is available on GitHub at: https://github.com/beards-lab/TriSeg-Digital-Twins.git.

**Figure 2:**
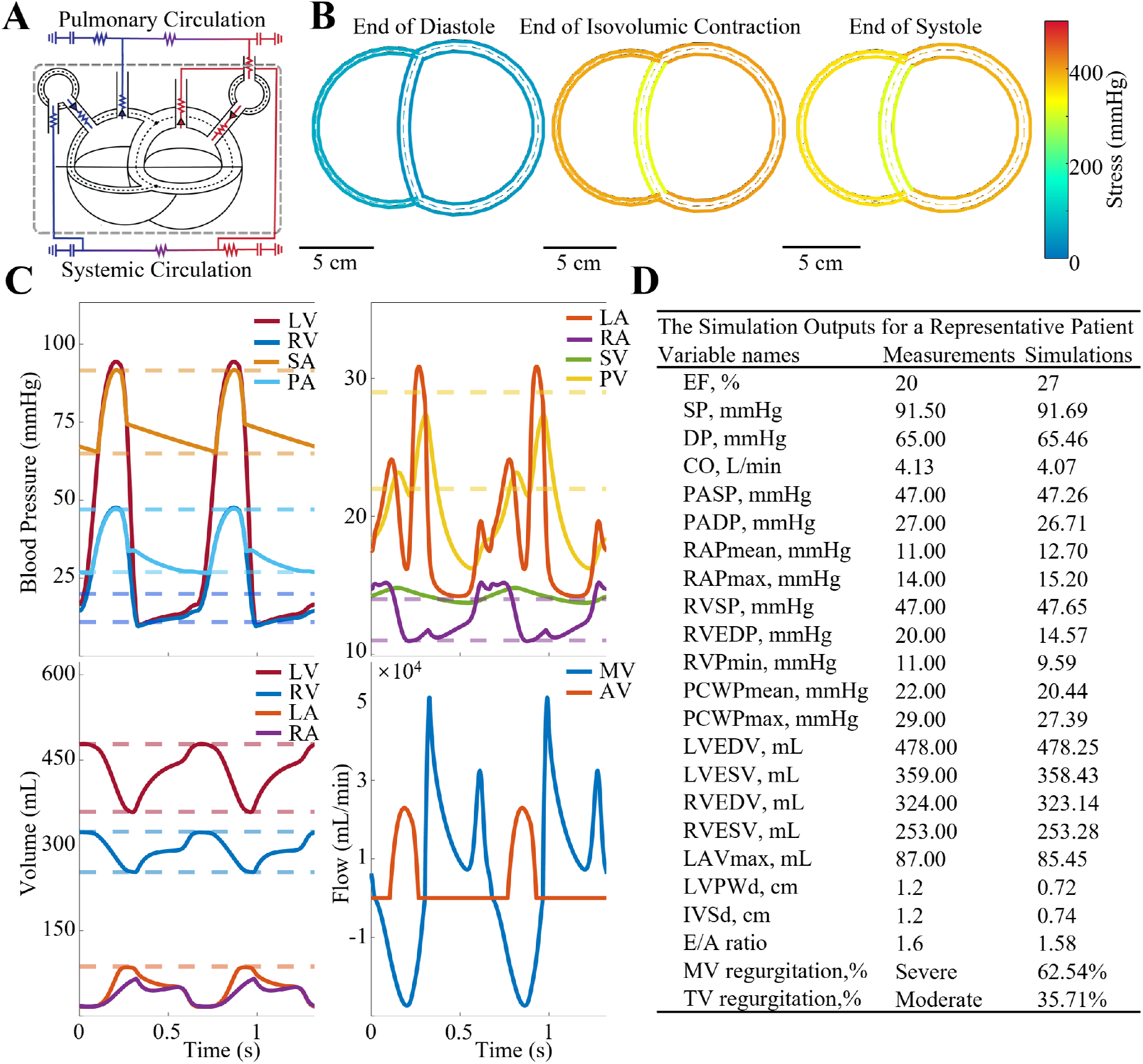
Digital twins identification. (A) The modified TriSeg heart model is integrated with a lumped parameter model to simulate closed-loop hemodynamics. (B) Model-predicted short-axis heart geometry is illustrated for a representative HFrEF patient at three phases during one cardiac cycle. The color represents myofiber stress in the wall. (C) Key simulation outputs are compared to data for the same patient. Dashed lines represent measurements from RHC, TTE, and CMR, while solid lines represent the simulation outputs. (D) The table compares model outputs to clinical measurements for the example patient. Valve regurgitation degree is reported as a categorical variable in the TTE reports and as regurgitation fraction in the simulations. TriSeg = three-wall segment, MV = mitral valve, AV = aortic valve. Other abbreviations are as previously defined at *Figure 1*.

### Unsupervised clustering

To determine the optimal number of phenogroups in the cohort, we employed K-means, hierarchical clustering, and Gaussian mixture model clustering, optimized using both the Davies-Bouldin index (DBI) and the gap statistic [21, 22]. After identifying the optimal number of clusters, we applied a hybrid approach combining K-means and hierarchical clustering to finalize the clustering results. A 3D volcano plot [23] was adapted to visualize differences across the three groups. Detailed methodologies are provided in the supplementary material *(Method S3)*.

### Prognostics AI model and validation

Overlapping features from the UMHS and UW cohorts, derived from clinical characteristics and digital twins, were used to train random survival forests (RSF), a non-linear machine learning method. The conventional Cox proportional hazards model was also developed. Both models were trained on the UMHS cohort data and externally validated on the UW cohort. The models were developed in R version 4.4.1., using the *randomForestSRC* and *glmnet* packages [24, 25], respectively. The predictive performance of the models was evaluated using the C-index, integrated area under the curve (iAUC), and time-dependent receiver operating characteristic (ROC) curves. To assess performance and quantify uncertainty, the RSF was trained 25 times with the same features and hyperparameters. Detailed methodologies are provided in the supplementary material *(Method S4)*.

### Statistical analysis

All statistical analyses were performed using GraphPad Prism 10.2.2. Continuous variables were analyzed via unpaired t-tests and one-way ANOVA. Categorical variables were compared using the chi-square test. Bonferroni correction was applied for multiple comparisons. Correlations were assessed using Pearson’s correlation coefficient. Statistical significance was defined as *p < 0.05, **p < 0.01, ***p < 0.001, and ****p < 0.0001.

## Results

### Digital twins yield novel patient-specific insights into cardiovascular state

We retrospectively reviewed 343 HF patients from the UMHS cohort, collecting data on 116 clinical characteristics. Among these patients, 215 were diagnosed with HFrEF, while 128 had a baseline left-ventricular ejection fraction (LVEF) estimated to be ≥ 50%. Although these patients are labeled as HFpEF for convenience, they represent a heterogeneous group, including patients with pulmonary arterial hypertension (N = 6) or hypertrophic cardiomyopathy (N = 32), contributing to a greater degree of heterogeneity compared to the general HFpEF population. The extracted baseline characteristics are summarized in Supplementary Table 1. Measured characteristics and paitent-specfic parameters from digital twins are provided in Supplementary Tables 2 & 3.

The identified digital twins represent a synthesis of anatomical measurements and volumetric flow data derived from TTE and CMR, along with pressure data from RHC. For example, simulations of a patient with HFrEF, a dilated left ventricle (LV), mitral insufficiency, and tricuspid insufficiency (Figure 2B, Supplementary Video 1) show cardiac anatomic and mechanical features consistent with the imaging and catheterization data for this individual. This individual’s simulation outputs, including pulmonary and systemic pressures, mitral and aortic valve flows, and ventricular and atrial volumes (Figure 2C) closely match the corresponding targets from clinical measurements (Figure 2D).

The relationships between simulated and measured variables for all subjects are illustrated in Supplementary Figure 1 and 2, indicating that digital twins robustly capture the diverse cardiovascular hemodynamic characteristics present in HF patients. Key parameter distributions, comparing HFpEF and HFrEF with normal subjects, are summarized in Supplementary Figure 3, revealing insights into contractility, power, stiffness, and vascular compliance across patient groups, which is consistent with HF pathophysiology.

### Digital twins reveal novel phenogroups representing distinct etiologies

Figure 3A and 3B show the 2D principle component projection of the space of functional parameters from digital twins and the corresponding PCA loadings. The PCA loadings reveal the parameter differences that underlie three phenogroups: Phenogroup 1 is characterized by patients with high vessel compliance and/or low vascular congestion; Phenogroup 2 is characterized by patients with high pulmonary resistance and elevated pericardial constraint; and Phenogroup 3 is characterized by patients with dilated and hypertrophic hearts, and elevated myocardial passive stiffness. Phenogroup 1 includes 61 HFrEF and 22 HFpEF patients; Phenogroup 2 includes 63 HFrEF and 88 HF-pEF patients; and Phenogroup 3 includes 91 HFrEF and 18 HFpEF patients (Figure 3C). There are no significant demographic differences across the phenogroups.

**Figure 3:**
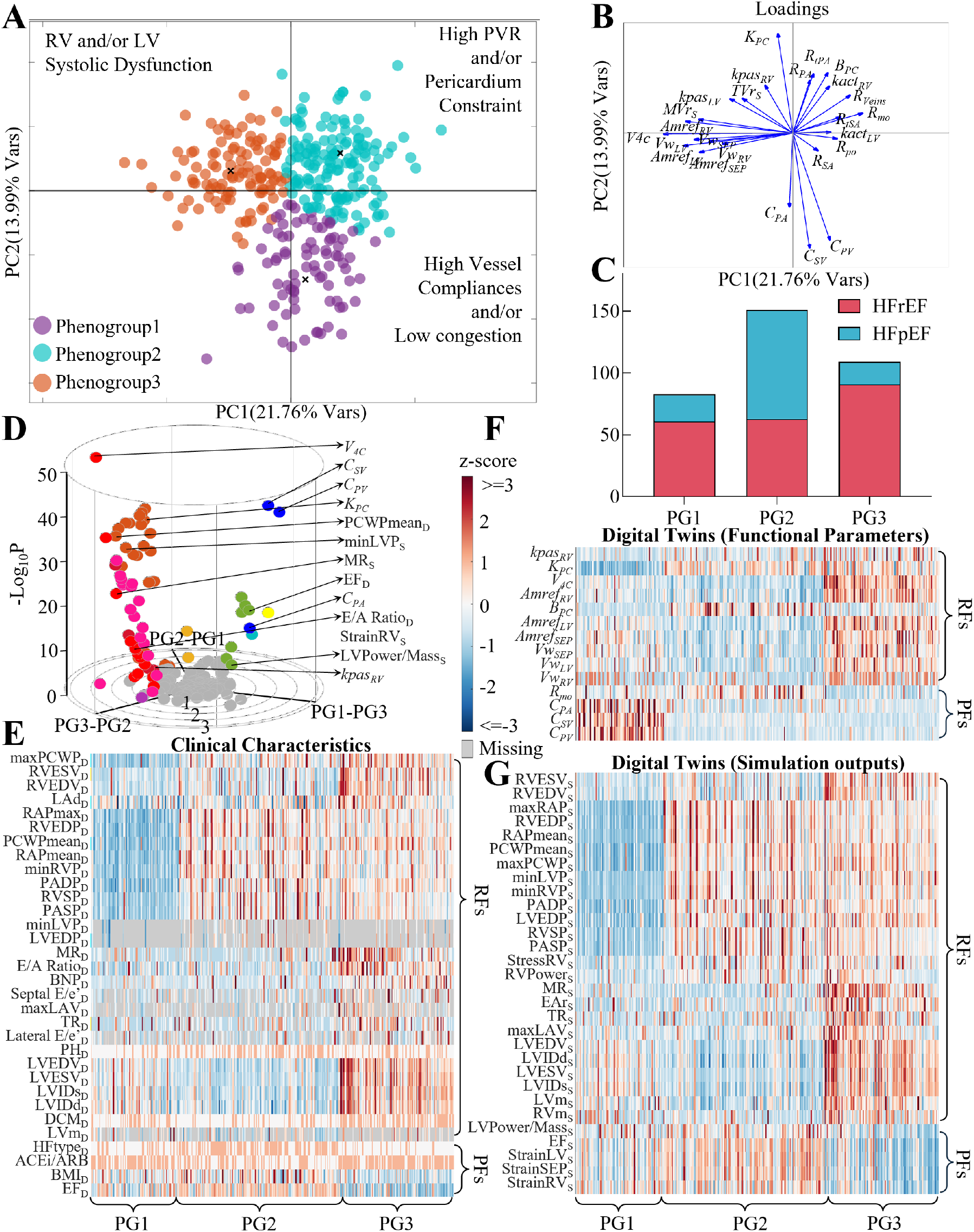
Precision phenotyping of heart failure patients. (A) Clustering results based on digital twins from UMHS cohort. (B) PCA loadings for PC1 and PC2. (C) Bar plot showing the distribution of diagnoses across phenogroups. (D) 3D volcano plot of clinical characteristics and digital twin features. (E- G) Heatmaps showing significant differences in clinical characteristics, functional parameters, and simulation outputs across phenogroups. PCA = principal component analysis, PC = principal component, PVR = pulmonary vascular resistance, PG = phenogroup, RF = risk factor, PF = protective factor. D = features from clinical data, S = features from simulation outputs. Other abbreviations are as defined in *Figure 1* and *Supplementary Table 3*.

A modified 3D volcano plot is used to visualize features that show significant differences between phenogroups (Figure 3D, Supplementary Video 2). Each dot in the plot represents a feature, where gray dots indicate insignificant differences, and colored dots indicate significant differences. Red indicates that variables progressively increase from phenogroups 1 to 2 to 3, while blue indicates that variables progressively decrease from Phenogroup 1 to 2 to 3. Features where Phenogroup 2 exhibits the highest value are colored orange with color gradient shifted towards red and green based on proximity to Phenogroups 1 and 3. Features where Phenogroup 2 exhibits the lowest value are colored purple, with color shifting towards pink and navy blue based on proximity to Phenogroups 1 and 3 (Supplementary Figure 4).

All significant features, including clinical characteristics and digital twins, are visualized using heatmaps (Figures 3E–G). Heatmap variables are listed based on graded changes across phenogroups, as captured by the 3D volcano plot. Variables are manually ordered, starting with those lowest in Phenogroup 1, followed by Phenogroup 2, and then Phenogroup 3. Consistent with the PCA loadings of the functional parameters (Figure 3A), Phenogroup 3 has the most extreme heart geometric parameters, associated with significant hypertrophy and dilation; Phenogroup 2 shows the highest pericardial constraint and mitral valve stenosis, and although pulmonary vascular resistance does not appear directly, it is reflected by the highest burden of PH. Phenogroup 1 exhibits similar heart geometry to Phenogroup 2 but has the highest vessel compliance and the lowest pericardial constraint (Figures 3E, F).

Hemodynamic and heart morphology measurements, along with corresponding simulation outputs, were assessed across phenogroups. These variables were categorized into two groups: functional and morphological abnormalities. Functional abnormalities were defined by elevated LV filling pressures (indicated by PCWP, LVEDP, E/A ratio and E/e’) and increased RV filling pressures (indicated by elevated RAP and RVEDP), as well as PH (evidenced by higher PASP, PADP, and RVSP). Morphological abnormalities are characterized by hypertrophy and heart enlargement, indicated by EF, LVEDV, LVESV, RVEDV, RVESV, LVIDd, LVIDs, IVSd, LVPWd, and LV and RV mass. Phenogroup 3 exhibits both functional and morphological abnormalities, while Phenogroup 2 shows functional abnormalities with generally normal heart morphology. In contrast, Phenogroup 1 has lower LV and RV filling pressures, lower pulmonary pressures, and relatively normal heart morphology (Figures 3E, G).

### Phenogroups identified by digital twins have distinct prognoses

Among the 343 patients in the UMHS cohort, over a 5-year follow-up period, 107 (31.2%) reached the primary composite endpoints. These included 94 all-cause deaths (27.4%), 16 LVAD implantations (4.7%), and 8 heart transplantations (2.3%) The freedom from the primary composite endpoint shows no significant difference between HFrEF and HFpEF (Figure 4A). However, there is a distinct pattern of association between the phenogroups identified by digital twins and the prognostic outcomes (Figure 4B, Supplementary Figure 5). Specifically, the survival probability within 5 years for the primary composite endpoint is highest in Phenogroup 1, followed by Phenogroups 2 and 3 (80.7%, 69.5%, and 58.7%, respectively) (Figure 5B). With regards to all-cause mortality, Phenogroup 1 also shows the highest survival probability (81.9%), whereas Phenogroups 2 and 3 have similar survival probabilities (70.9% and 67.9%), resulting in a non-significant difference across the three phenogroups (Figure 4C). The *MAGGIC* score, widely used for predicting all-cause mortality in HF patients [26], was also calculated for these three phenogroups. Phenogroup 3 has the highest MAGGIC score, while Phenogroups 1 and 2 have comparable scores, which is partially consistent with the Kaplan-Meier survival differences (Figure 4D).

**Figure 4:**
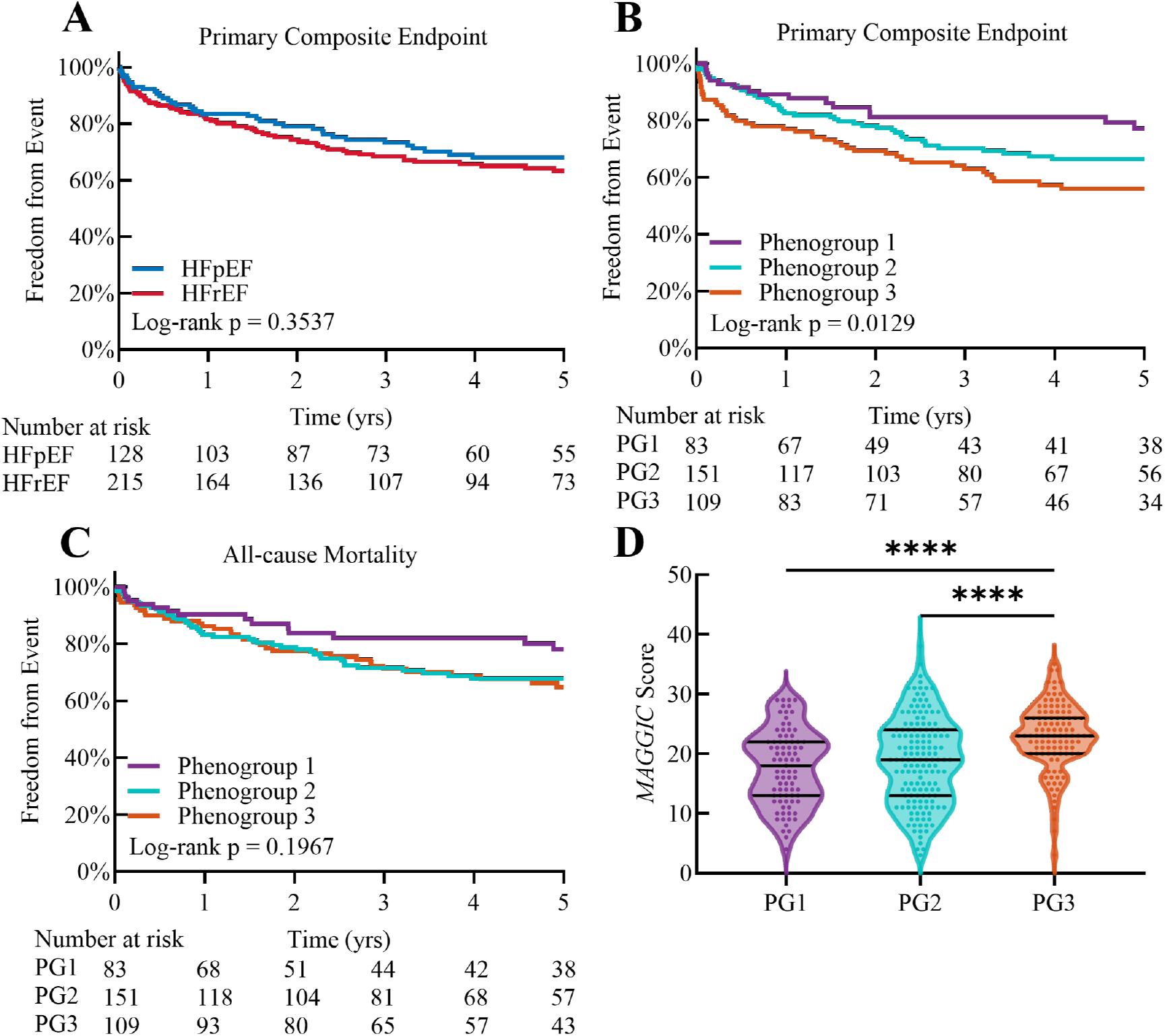
Differences in prognostic outcomes across phenogroups. (A) Kaplan-Meier plot for the primary composite endpoint in the UMHS cohort, stratified by diagnosis. Kaplan-Meier plot for the (B) primary composite endpoint and (C) all-cause mortality in the UMHS cohort, stratified by phenogroups. (D) Differences in *MAGGIC* scores across phenogroups. The primary composite endpoint includes all-cause mortality, LVAD implantation, and heart transplantation. Abbreviations are as defined above.

**Figure 5:**
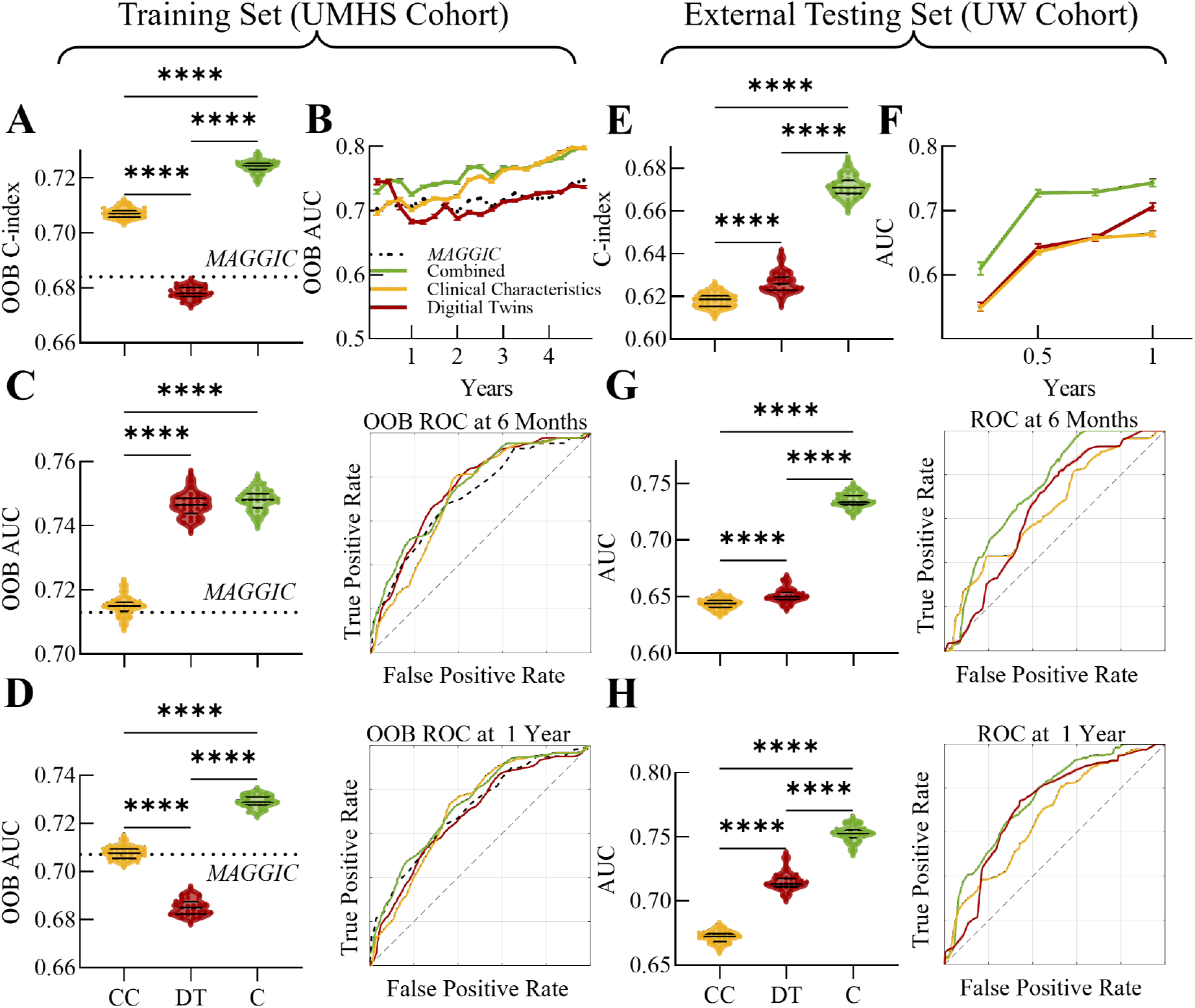
Digital twins improve prognostic AI performance. The prognostic AI, referred to here as the RSF model, is trained using clinical characteristics, digital twins, and their combination from the UMHS cohort. It is evaluated using (A) OOB C-index and (B) time-dependent AUC. The time-dependent ROC curves and corresponding AUCs at (C) 6 months and (D) 1 year are shown. The AI’s performance is also externally validated using the UW cohort, with evaluations based on (E) the C-index and (F) time-dependent AUC. Time-dependent ROC curves and corresponding AUCs at (G) 6 months and (H) 1 year are also displayed. Data are presented as violin plots or mean ± SD. RSF = random survival forests, OOB = out-of-bag, AUC = area under the curve, ROC = receiver operating characteristic, CC = clinical characteristics, DT = digital twins, C = combination of CC and DT.

The multivariable Cox proportional hazards model was employed to assess the association between phenogroups and clinical outcomes using Phenogroup 1 as the reference group. Well-established HF prognostic factors were selected as covariates [26], while those involved in the digital twin identification process were excluded to avoid confounding effects. Phenogroup 3 is significantly associated with a higher risk of the primary composite endpoint [hazard ratio (HR) 2.72, 95% confidence interval (CI) 1.53-5.06] and all-cause mortality (HR 1.98, 95% CI 1.08-3.80)(Table 1). Phenogroup 2 initially appears to be associated with higher risk of the primary composite endpoints and all-cause mortality, as it shows higher rates for both primary composite endpoints (30.5% vs. 19.3%) and all-cause mortality (29.1% vs. 18.1%) compared to Phenogroup 1. However, after adjustments for covariates, these associations are not statistically significant (Table 1).

**Table 1.** Association between phenogroup membership and the risk of different adverse clinical outcomes using a multivariable Cox proportional hazard analysis.

|  | Phenogroup 1 (*n* = 83) |  | Phenogroup 2 (*n* = 151) |  | *P* | Phenogroup 3 (*n* = 109) |  | *P* |
| --- | --- | --- | --- | --- | --- | --- | --- | --- |
|  | Events | Reference | Events | HR (95% CI) |  | Events | HR (95% CI) |  |
| Primary composite endpoint<sup>a</sup> | 16 (19.3%) | Ref. | 46 (30.5%) | 1.11 (0.63–2.06) | 0.73 | 45 (41.3%) | 2.72 (1.53–5.06) | 0.001 |
| All-cause mortality | 15 (18.1%) | Ref. | 44 (29.1%) | 1.14 (0.63–2.16) | 0.68 | 35 (32.1%) | 1.98 (1.08–3.8) | 0.03 |

CI, confidence interval; HR, hazard ratio.
Outcomes were adjusted for age, sex, smoking status, race, history of diabetes, hypertension, chronic obstructive pulmonary disease (COPD), baseline creatinine levels, usage of angiotensin-converting enzyme inhibitors (ACEi), angiotensin II receptor blockers (ARBs), and beta blockers, as well as for whether the patient had a first diagnosis of heart failure within the past 18 months.
<sup>a</sup> A composite endpoint includes all-cause mortality, LVAD implantation, or heart transplantation

To interpret the differences phenogroup outcomes (Figures 4B, C), we identified risk and protective factors among features, based on whether or not they are increased or decreased in value or frequency in Phenogroup 1 compared to 3 (Figures 3E, F, and G). Most of the risk and protective factors from clinical characteristics are well-established in relation to HF prognosis. Risk factors include elevated leftsided filling pressures, right-sided filling pressures, BNP levels, abnormal heart geometry, as well as PH and atrioventricular valve insufficiencies. Protective factors include higher BMI, better EF, and more frequent use of ACEi and ARBs (Figure 3E). Beyond these clinical characteristics, the functional parameters of digital twins identify additional risk and protective factors, some of which are difficult or impossible to measure clinically. Risk factors include increased RV passive stiffness and pericardial constraint, while protective factors include preserved pulmonary and systemic vascular compliance and mitral valve resistance (Figure 3F). Simulations from digital twins show substantial overlap with clinical risk factors and also identify novel risk factors like RV power output and RV free wall maximum stress, along with protective factors such as the LV power/mass ratio and maximum strain of the LV, RV, and septum (Figure 3G).

### Digital twins enhancing predictive AI performance

We further extended our analysis by using digital twin features in a prognostic ML model for predicting primary composite endpoints. The RSF predictive model, selected based on its performance [16, 18], is trained using features present in both the UMHS and UW cohorts, using features from clinical characteristics only, digital twins only, and a combination of both from UMHS cohort. The selected features for the RSF input are shown in Table 2, ranked by variable importance.

**Table 2.** Selected variables for random survival forest.

| Clinical Characteristics<br>(*n* = 15) | Digital Twins<br>(*n* = 20) | Combined<br>(*n* = 21) | Extended *MAGGIC* Score<br>(*n* = 5) |
| --- | --- | --- | --- |
| Age | *Compliance of Systemic Veins* | Age | *MAGGIC* score |
| Hemoglobin | *Compliance of Pulmonary Veins* | Hemoglobin | *E/A Ratio* |
| Creatinine | *Constant Pericardium Constraint* | Creatinine | **LV Inner Diameter (end systole)** |
| BNP | **RVEDP** | Cardiac Output | **Mean Mitral Valve Pressure Gradient** |
| Max RA Pressure | *Resistance of Veins* | LV Posterior Wall Thickness | *Constant Pericardium Constraint* |
| Cardiac Output | **Tricuspid Valve Regurgitation Fraction** | Interventricular Septum Thickness |  |
| LVESV | *Resistance of Pulmonary Valve* | E/A ratio |  |
| LV Posterior Wall Thickness | *Resistance of Mitral Valve* | *Compliance of Systemic Veins* |  |
| E/A Ratio | **LV Inner Diameter (end diastole)** | *Resistance of Mitral Valve* |  |
| LVEDV | **LVEDP** | **Mean Mitral Valve Pressure Gradient** |  |
| LV Inner Diameter (end diastole) | *Resistance of Tricuspid Valve (backwards flow)* | **Peak Pulmonary Valve Pressure Gradient** |  |
| <u>Diabetes Mellitus</u> | **Mean Mitral Valve Pressure Gradient** | *Pulmonary Vascular Resistance* |  |
| RVSP | **Peak Pulmonary Valve Pressure Gradient** | <u>End Stage Renal Disease</u> |  |
| End Stage Renal Disease | **E/A ratio** | <u>Diabetes Mellitus</u> |  |
| PASP | *Pulmonary Vascular Resistance* | *Resistance of Pulmonary Valve* |  |
|  | **PADP** | **Maximum Stress of Septum** |  |
|  | **DBP** | NYHA Class |  |
|  | *Exponential Pericardium Constraint* | RVEDV |  |
|  | **LVESV** | **Maximum Strain of LV** |  |
|  | *Transmural Resistance of Systemic Arteries* | LV inner diameter (end diastole) |  |
|  |  | Minimum RV pressure |  |

Italics represents functional parameters from digital twins, bold represents simulation outputs from digital twins, and underlined represents comorbidities. Normal text without any formatting represents other clinical characteristics. Abbreviations are as defined above.

The out-of-bag (OOB) C-index of the RSF model based on combined features is the highest (0.724), compared to models using only clinical characteristics (0.707) or digital twins (0.678). For reference, the C-index for the *MAGGIC* score is 0.684 (Figure 5A). Time-dependent ROC analysis shows that the RSF model based on combined features has the highest predictive power over the entire time range, with an OOB iAUC of 0.744, compared to 0.721 for clinical characteristics and 0.719 for digital twins (Figure 5B). Interestingly, the RSF based on digital twins demonstrates the highest predictive power within the first year, with OOB AUCs for clinical characteristics, digital twins, and combined features of 0.715, 0.747, and 0.748, respectively, at six months (Figure 5C), and 0.708, 0.685, and 0.729, respectively, at one year (Figure 5D), indicating that the information from digital twins enhances short-term predictions.

These findings are externally validated using the UW cohort. Since in the UW cohort of 86 patients only two experienced death and none underwent LVAD implantation or heart transplantation, we used the trained RSF models to predict the composite endpoint of all-cause mortality and rehospitalization. The external validation confirms that the RSF model based on combined features has the highest Cindex (0.671), outperforming models using either clinical characteristics (0.618) or digital twins (0.626). Compared to the training set, RSF models based on combined features or digital twins maintain relatively stable predictive power, while the model based solely on clinical characteristics shows a bigger decrease in predictive power for the validation cohort compared to the training data (Figure 5E). The time-dependent ROC results are consistent with these conclusions, with iAUCs of 0.613,0.623, and 0.690, respectively (Figure 5E). In terms of 6-month and 1-year predictions, the RSF models based on combined features shows the highest performance, with AUCs of 0.735 at six months and 0.753 at one year. The predictive power for the digital twins-based model surpasses that of the clinical features-based model (Figures 5G, H). We also validated the model using only HFpEF patients. While the combined model continued to show the highest performance, the overall performance decreases for the HFpEF only validation group (*n* = 32, Supplementary Figure 6). Additionally, in the UW cohort, two HFpEF patients who reached the allcause mortality endpoint consistently had higher risk scores than the surviving HFpEF patients, regardless of whether predictions were based on clinical features, digital twins, or the combination (Supplementary Figure 7).

We also built RSF models using the *MAGGIC* score combined with additional features from digital twins (Extended *MAGGIC* score) based on UMHS cohort. By adding four features from digital twins (Table 2), the OOB C-index increased from 0.684 to 0.731 (Figure 6A). The model’s predictive power remained stable throughout the followup period, with the iAUC improving from 0.710 to 0.768 (Figure 6B). At six months, the AUC for the RSF model incorporating additional features reached 0.788, compared to 0.713 for the *MAGGIC* score alone (Figure 6C), and at one year, these values were 0.756 and 0.707, respectively (Figure 6D). Among all tested RSF models, the extended *MAGGIC* score achieved the best OOB performance (Figures 5,6), suggesting a mix of linear and non-linear relationships between the predictors and outcomes.

**Figure 6:**
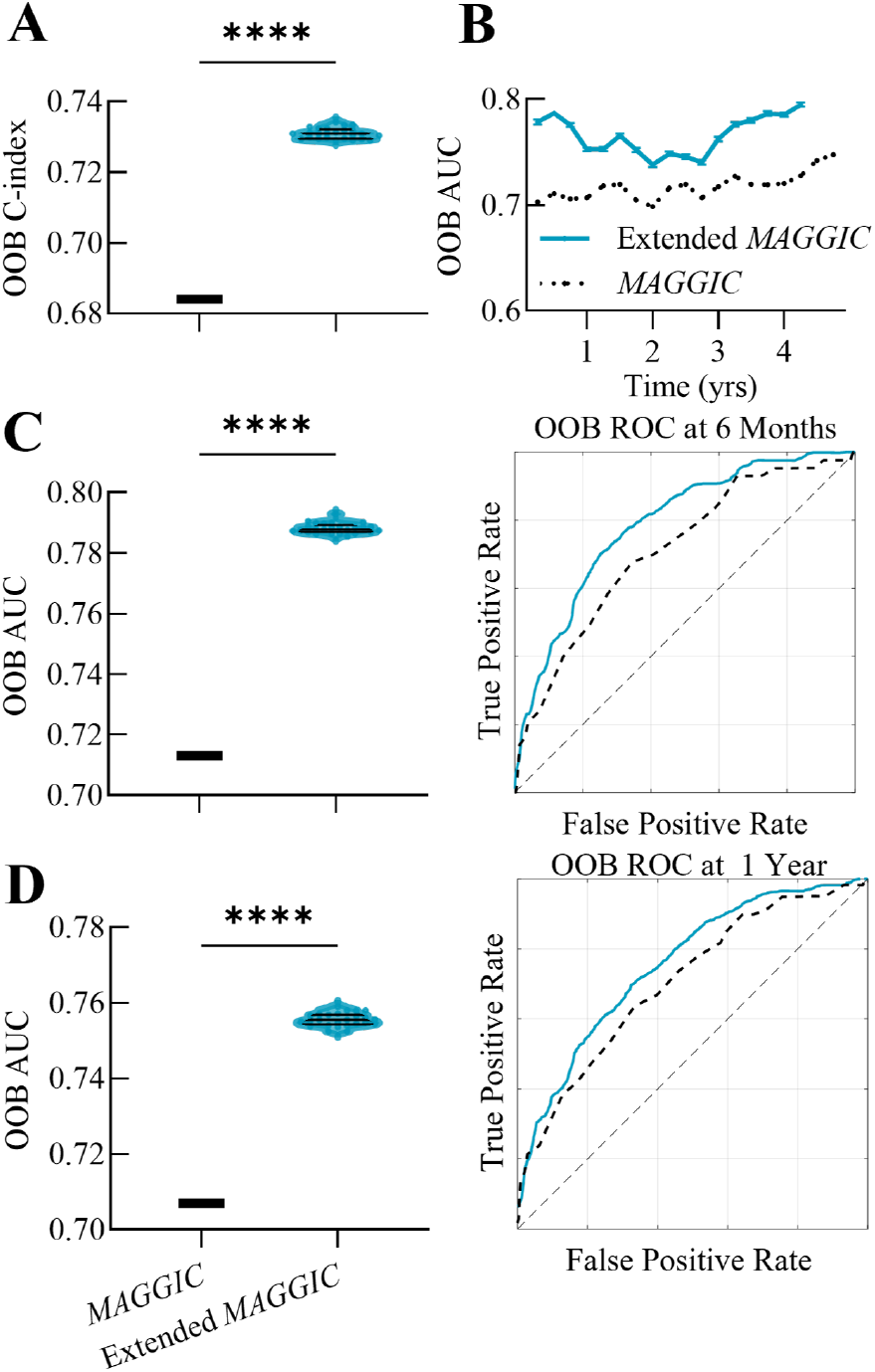
Extended *MAGGIC* score. The Extended *MAGGIC* score combines the traditional *MAGGIC* score with additional features derived from digital twins. Differences in (A) the OOB C-index and (B) time-dependent AUC between the Extended *MAGGIC* score and the original *MAGGIC* score are illustrated. Time-dependent ROC curves at (C) 6 months and (D) 1 year, along with the corresponding AUCs, are also shown. Data are presented as violin plots or mean ± standard deviation from the UMHS cohort. Abbreviations are as defined above.

## Discussion

### Summary

The present study introduces a novel ML approach to assess patients with HF, using physiology-informed patientspecific simulations of cardiovascular function. Digital twins not only provide individualized mechanistic insights but also, through unsupervised ML, reveal three distinct phenogroups linked to differential clinical outcomes. This approach yields interpretable insights into the underlying mechanisms of these phenogroups: Phenogroup 3 includes patients with severe RV and/or LV systolic dysfunction; Phenogroup 2 consists of patients with high pulmonary vascular resistance or significant pericardial constraint; and Phenogroup 1 comprises patients with high vascular compliance and/or low congestion. Functional differences across these phenogroups reveal baseline risk and protective factors. Furthermore, results from supervised ML suggest that digital twins can complement and improve the performance of prognostic AI models.

This simulation-based approach offers several additional advantages. It can correct inconsistencies and inaccuracies in clinical measurements, and can be used to impute missing data (Figures 4E, G). It estimates functional parameters that are otherwise impossible to measure directly, providing a more complete and accurate representation of the patient’s disease state than raw clinical data alone. In addition, the digital twin framework has the potential to empower simulations of patient-specific prognoses and therapy outcomes. Ultimately, this approach has the potential to enhance the ability to focus on the most critical aspects of a patient’s condition, leading to individualized care and management.

### Phenomapping

Phenomapping in HF, using ML techniques to identify distinct subgroups among HF patients [8–15, 19], faces several key limitations. The data sources are often chosen based on ease of access and prior knowledge, leading to significant variability across studies. Phenogroups identified in different studies often differ substantially in both number and composition. While certain subgroups may share some clinical characteristics across studies, the classification boundaries are often unclear, with considerable overlap between groups in different studies [27]. Additionally, the heterogeneity of HF as a syndrome is usually overlooked, and studies tend to under-explore important mimickers, such as amyloidosis, which can obscure the phenomapping results [28]. Moreover, many studies lack hemodynamic measurements, which are crucial, especially in the early stages of HFpEF. These challenges highlight the need for more comprehensive data integration, the inclusion of under-explored variables like hemodynamics, and a focus on discovering novel insights beyond existing clinical knowledge.

Our digital twin hybrid with unsupervised ML helps to overcome some of these limitations. Here phenomapping is driven by simulations matched to hemodynamic and heart geometrical measurements. Moreover, our approach avoids using dimensionality reduction methods, which, while powerful, often result in less interpretable outcomes. Instead, our computational model maps patient data onto physiological functional parameters. Because physiological knowledge is embedded in the simulation framework, it yields identification of novel phenogroups and risk factors, providing more interpretable insights than those available from purely datadriven approaches.

### Prognostic value

Since the phenotypes of patients represented in digital twins encapsulate physiological/pathological interrelationships among clinical data, the digital twin is a robust vehicle for incorporating domain knowledge into supervised ML. Integration of digital twin features into AI-based analyses of patient data improves survival prediction and generalizability of predictive models.

Our analysis identifies novel risk and protective factors, as well as predictors for outcomes in patients with HF. Factors derived from clinical characteristics, such as high BNP levels being associated with poor outcomes, align with prior knowledge and are not particularly novel. However, the factors identified from digital twins, which are often difficult or impossible to measure directly, represent promising new insights. First, both supervised and unsupervised ML revealed that high vascular compliance is associated with better outcomes, a potential prognostic factor that has not been well investigated [29]. Second, passive stiffness of the ventricle emerged as a risk factor from unsupervised ML. While this factor cannot be measured directly, it can be assessed through indicators of diastolic dysfunction and is increasingly recognized for its prognostic value in both HFrEF and HFpEF [30]. Additionally, pericardial constraint was identified as a risk factor by both supervised and unsupervised ML. While researchers have recognized the potential role of the pericardium in the etiology of HFpEF [31], its significance in prognosis has been underexplored and warrants further investigation. Lastly, some known but challenging-to-measure risk factors, such as LVEDP, were also identifiable through digital twins. This capability highlights the substantial application potential of digital twins, suggesting they can provide critical insights that are difficult to derive from raw clinical data alone.

## Data Availability

All data produced are available online at https://github.com/beards-lab/TriSeg-Digital-Twins.git

