## Supplemental Material for "Identification of Digital Twins to Guide Interpretable AI for Diagnosis and Prognosis in Heart Failure"

### 1. Figures and Tables

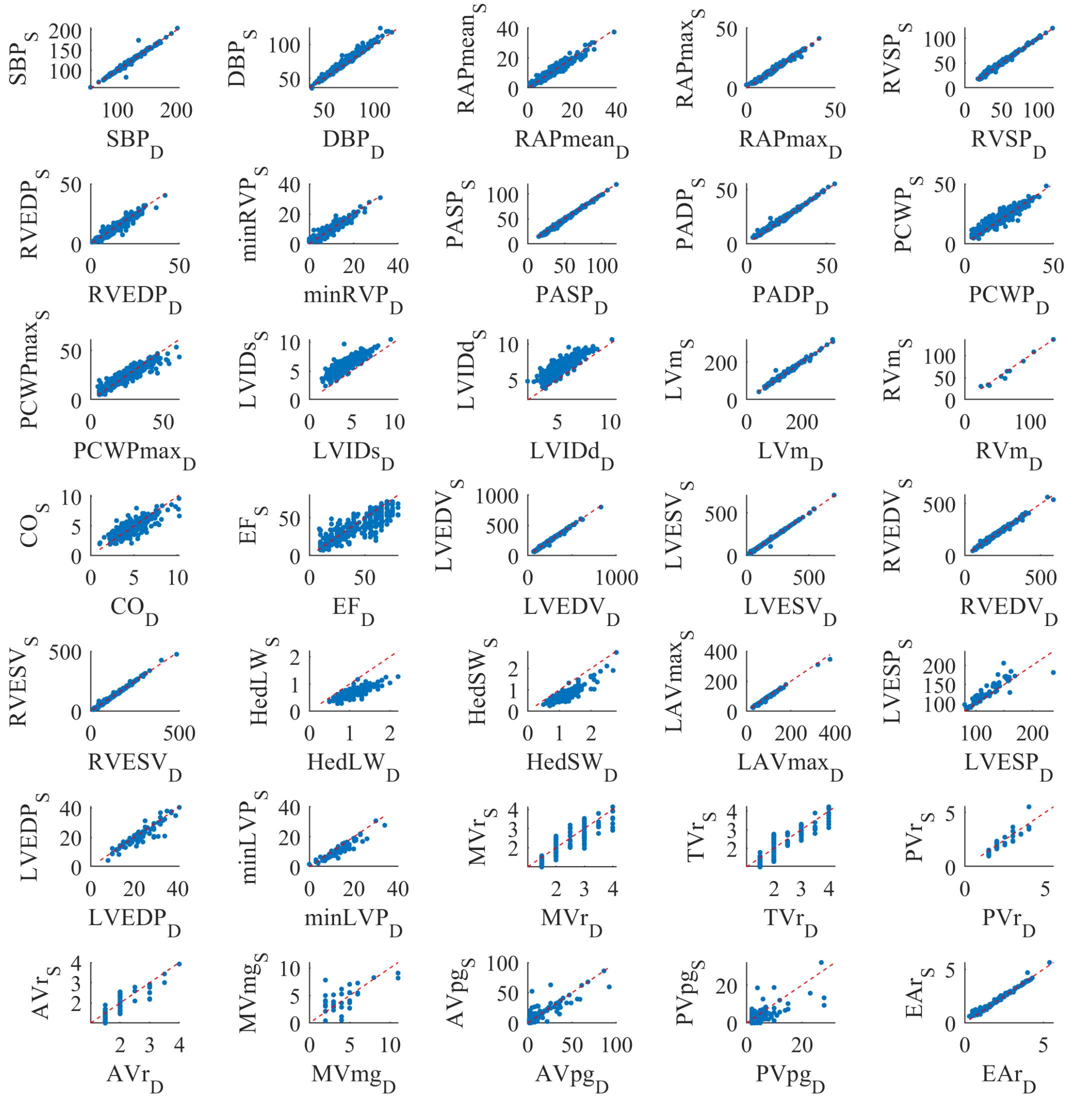


***Supplementary Figure* 1: Comparison of clinical measurement targets to and simulation outputs for variables used for model identification**

Each panel presents a scatter plot comparing clinical measurement targets with corresponding simulation outputs for the complete UMHS cohort of 343 patients.

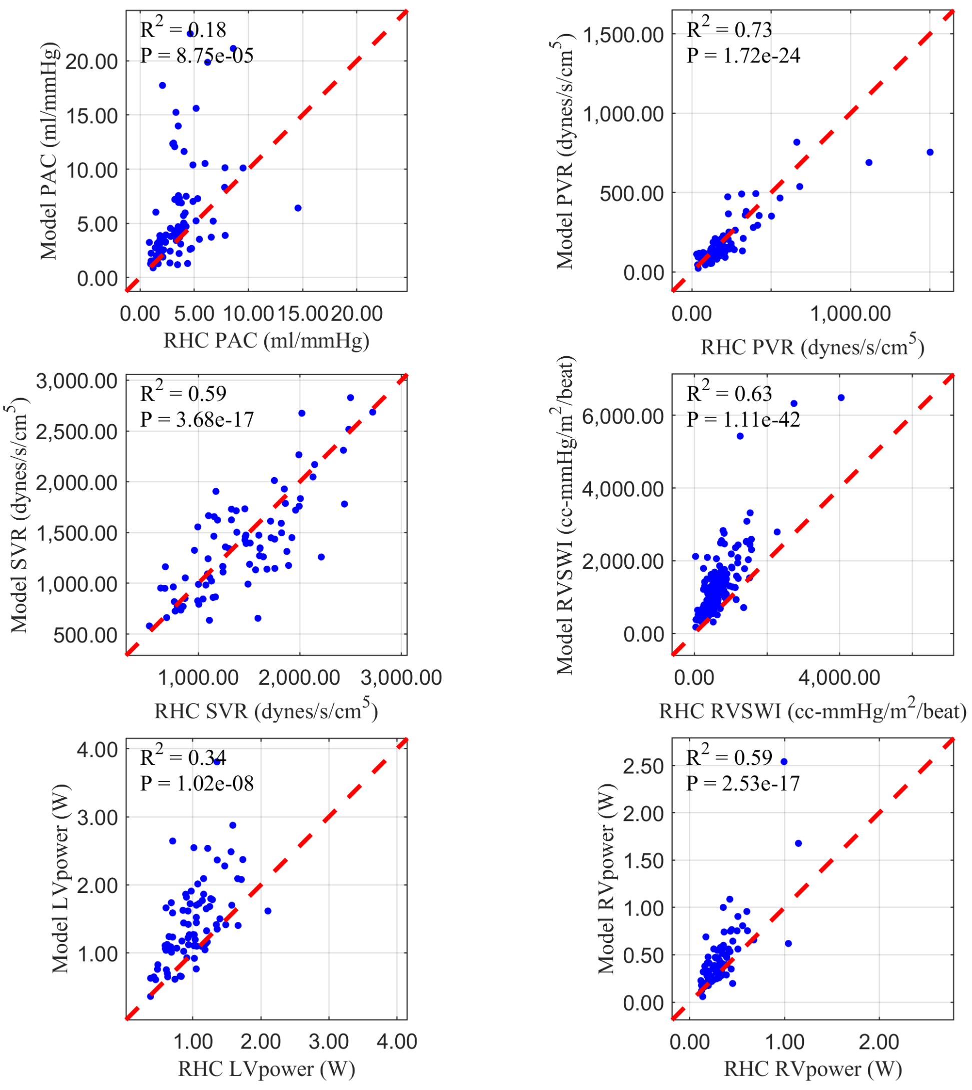


***Supplementary Figure* 2: Comparison of RHC measurements and corresponding simulation outputs**
PAC = Pulmonary Arterial Compliance, PVR = Pulmonary Vascular Resistance, SVR = Systemic Vascular Resistance, RVSWI = Right Ventricular Stroke Work Index

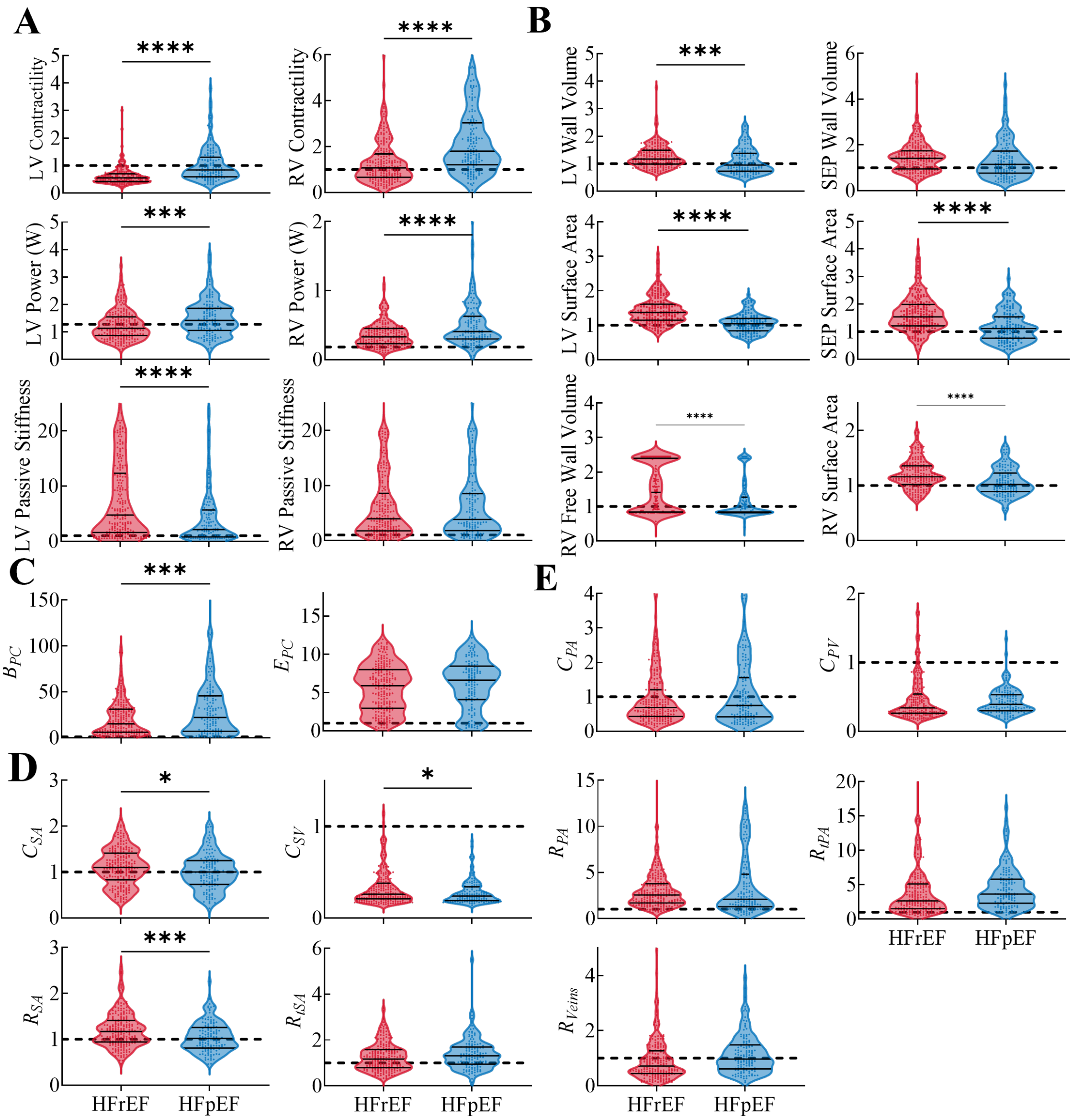


***Supplementary Figure* 3: Distributions of model parameter values for HFrEF and HFpEF groups**

The data are presented as violin plots with medians and interquartile ranges. Parameter values are reported relative to control representations of baseline healthy male and female. Each dot represents an individual patient from the UMHS cohort, with red indicating HFrEF and blue indicating HFpEF. Functional parameters are organized as: heart mechanical properties (A), heart geometry (B), pericardium constraint (C), systemic circulation (D), and pulmonary circulation (E). HFrEF = heart failure with reduced ejection fraction, HFpEF = heart failure with preserved ejection fraction. Other abbreviations are defined in Supplementary Table 3. *p < 0.05, **p < 0.01, ***p < 0.001, ****p < 0.0001.

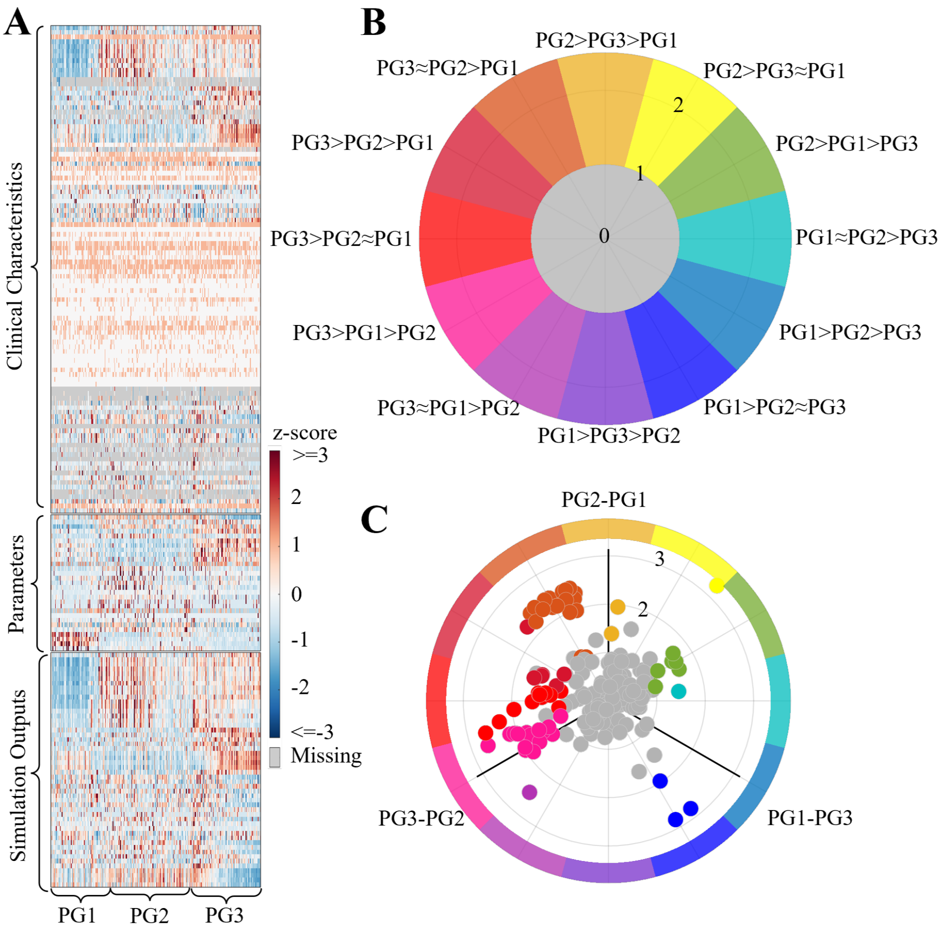


***Supplementary Figure* 4: 3D volcano plot design**

(A) Heatmap of all clinical characteristics and digital twin features, sorted by phenogroups. (B) The base of the 3D volcano plot is structured as a circular polar coordinate system, where the polar axis represents differences in three distinct directions. Colored regions highlight features with significant differences (z-score difference > 1), with each color indicating a specific type of difference among the three phenogroups. Gray denotes non-significant differences. (C) After integrating differences with p-values along the z-axis, the final significantly different features presented in Figure 3 are also plotted on the polar axis.

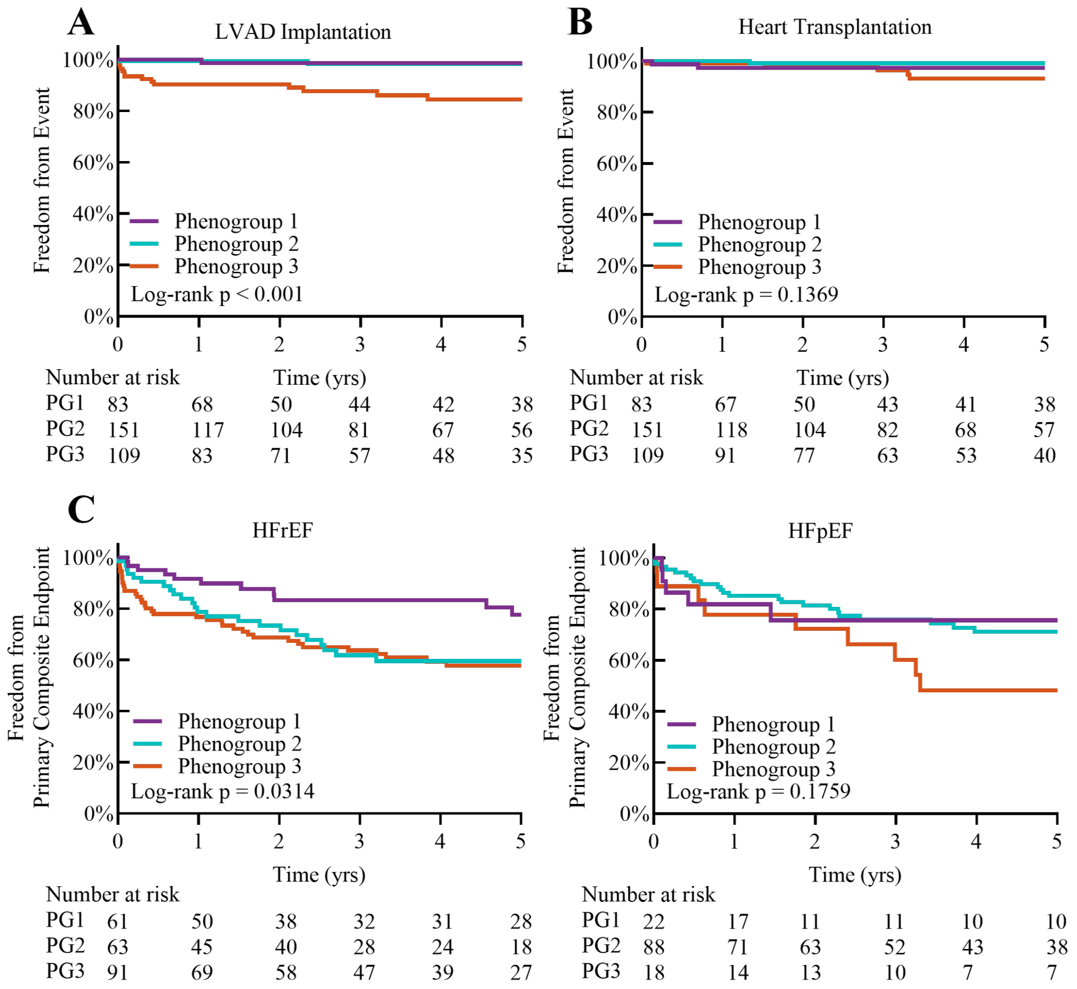


***Supplementary Figure* 5: Kaplan-Meier plots for (A) LVAD implantation and (B) heart transplantation, stratified by phenogroups within the UMHS cohort, and (C) the primary composite endpoint, stratified by HFrEF and HFpEF subgroups and further stratified by phenogroups within the UMHS cohort.**

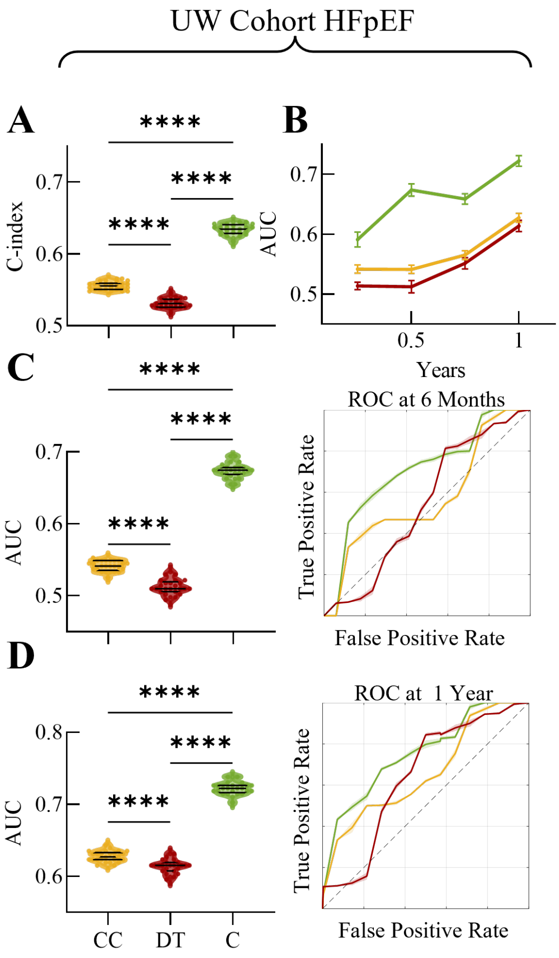


***Supplementary Figure* 6: RSF valadation by only patients with HFpEF**

The RSF's performance is also externally validated using only HFpEF patients from UW cohort, with evaluations based on (A) the C-index and (B) time-dependent AUC. Time-dependent ROC curves and corresponding AUCs at (C) 6 months and (D) 1 year are also displayed.

Abbreviations are defined in Figure 5.

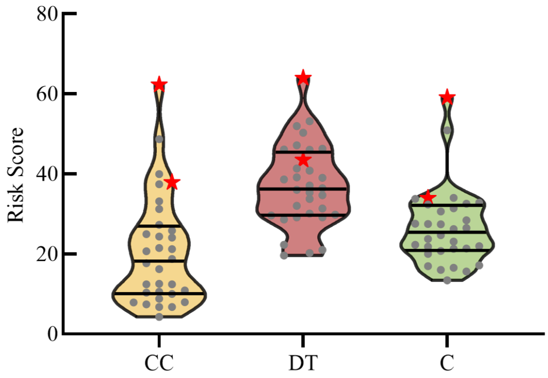


***Supplementary Figure* 7: Distribution of RSF risk scores in HFpEF patients within the UM cohort.**
The RSF models—whether based on clinical features, digital twins, or a combination—demonstrate that deceased subjects have higher risk scores compared to survivors in patients with HFpEF, with the largest difference observed in the combined group.

**Supplementary Table 1**
#### Baseline and clinical characteristics of the study participants in the UMHS cohort

| | HFpEF (n=128) | HFrEF (n=215) |
|---|---|---|
| **Age, years** | 57.3 ± 14.4 | 53.7 ± 16.1 |
| **Male, n (%)** | 66 (51.5) | 120 (55.8) |
| **Race, n (%)** | | |
| White | 95 (74.2) | 155 (72.1) |
| Black | 21 (16.4) | 46 (21.4) |
| Other | 12 (9.4) | 14 (6.5) |
| **NYHA functional class, n (%)** | | |
| I | 15 (11.7) | 19 (8.8) |
| II | 39 (30.5) | 83 (38.6) |
| III | 61 (50.8) | 92 (42.8) |
| IV | 13 (10.2) | 21 (9.8) |
| **Newly diagnosed HF within 18 months, n (%)** | 77 (60.2) | 167 (77.7) |
| **Comorbidities, n (%)** | | |
| Coronary artery disease | 49 (38.3) | 98 (45.6) |
| Hypertension | 93 (73.7) | 142 (66.0) |
| Hyperlipidemia | 73 (57.0) | 115 (53.5) |
| Valvular disease | 49 (38.3) | 91 (42.3) |
| Hypertrophic cardiomyopathy | 32 (25.0) | 10 (4.7) |
| Dilated cardiomyopathy | 6 (4.7) | 124 (57.7) |
| Amyloidosis | 10 (7.8) | 0 (0.0) |
| Atrial fibrillation | 36 (28.1) | 41 (19.1) |
| Other type arrhythmia | 49 (38.3) | 93 (43.3) |
| Diabetes mellitus | 38 (30.0) | 60 (27.9) |
| Chronic obstructive pulmonary disease | 26 (20.3) | 32 (14.9) |
| Pulmonary hypertension | 63 (49.2) | 58 (27.0) |
| Connective tissue disease | 17 (13.3) | 19 (8.8) |
| End-stage renal disease | 41 (18.8) | 79 (36.4) |
| Respiratory failure | 36 (28.1) | 51 (23.7) |
| Aortic aneurysm/dissection | 13 (10.2) | 14 (6.5) |
| Pericardial disease | 20 (15.6) | 10 (4.7) |
| Hyperthyroidism | 1 (0.8) | 2 (0.93) |
| Hypothyroidism | 18 (14.1) | 32 (14.9) |
| **Clinical characteristic and laboratory data** | | |
| Heart rate, bpm | 74.5 ± 15.2 | 79.7 ± 15.4 |
| Ejection fraction, % | 63.4 ± 8.2 | 16.1 ± 1.1 |
| Systolic blood pressure, mmHg | 120.4 ± 20.5 | 112.6 ± 22.8 |
| Diastolic blood pressure, mmHg | 69.5 ± 11.1 | 71 ± 14.1 |
| Cardiac output, L/min | 5.0 ± 1.6 | 4.4 ± 1.4 |
| Hemoglobin, g/dL | 12.4 ± 2.4 | 12.6 ± 2.5 |
| Fasting glucose | 100.9 ± 37.6 | 122 ± 54.5 |
| Body mass index, kg/m$^2$ | 30.6 ± 7.9 | 28.9 ± 7.2 |
| Serum creatine, umol/L | 106.9 ± 69.7 | 108.3 ± 62.3 |
| B-type natriuretic peptide, pg/mL | 564.5 ± 913.7 | 829.6 ± 953.9 |
| Thyroid-Stimulating Hormone, mIU/L | 2.8 ± 3.7 | 3.7 ± 5.3 |
| **Medication and Device Therapy, n (%)** | | |
| ACE inhibitor | 27 (21.1) | 100 (46.5) |
| ARB or ARNi | 15 (11.7) | 65 (30.2) |
| $\beta$-Blocker | 72 (56.3) | 170 (79.1) |
| Diuretics | 90 (70.3) | 184 (85.6) |
| Endothelin receptor antagonists | 2 (1.6) | 1 (0.5) |
| Calcium channel blocker | 49 (38.3) | 66 (30.7) |
| Nitrates | 27 (21.1) | 92 (42.8) |
| Aspirin | 88 (68.8) | 164 (76.3) |
| Statin | 65 (50.8) | 128 (59.5) |
| SGLT2 inhibitor | 7 (5.5) | 20 (9.3) |
| Implantable Cardioverter-Defibrillator | 12 (9.4) | 40 (18.6) |
| Cardiac Resynchronization Therapy | 0 (0.0) | 8 (3.7) |
| Pacemaker | 9 (7.0) | 7 (3.3) |
| O2 supply | 31 (24.2) | 48 (22.3) |
| **Social History, n (%)** | | |
| Current Smoker | 15 (11.7) | 40 (18.6) |
| Alcohol abuse | 65 (50.8) | 95 (44.2) |
| Drug abuse | 11 (8.6) | 17 (7.9) |
| ***MAGGIC* Risk Score** | 17.7 ± 7.0 | 21.0 ± 6.5 |

Data are presented as mean ± standard deviation for continuous variables or *n* (%) for categorical variables.
SGLT2, Sodium-glucose cotransporter-2.

**Supplementary Table 2**
#### Clinical characteristics used for the identification of digital twins

| | HFpEF (n=128) | HFrEF (n=215) |
|---|---|---|
| **Height, cm** | 170.2 ± 11.0 | 171.0 ± 10.7 |
| **Weight, kg** | 88.4 ± 24.6 | 84.6 ± 23.3 |
| **Hemodynamic measurements** | | |
| Heart rate, bpm | 74.5 ± 15.2 | 79.7 ± 15.4 |
| Systolic blood pressure, mmHg | 120.4 ± 20.5 | 112.6 ± 22.8 |
| Diastolic blood pressure, mmHg | 69.5 ± 11.1 | 71.0 ± 14.6 |
| Cardiac output, L/min | 5.0 ± 1.6 | 4.4 ± 1.4 |
| Pulmonary systolic blood pressure, mmHg | 49.8 ± 21.0 | 42 ± 14.1 |
| Pulmonary diastolic blood pressure, mmHg | 24.1 ± 9.6 | 22 ± 8.7 |
| Right atrial mean pressure, mmHg | 13.3 ± 6.5 | 11.1 ± 5.9 |
| Right atrial maximum pressure, mmHg | 16.6 ± 6.9 | 14.2 ± 6.6 |
| Right ventricle end systolic pressure, mmHg | 50.6 ± 20.7 | 42.4 ± 14.2 |
| Right ventricle end diastolic pressure, mmHg | 15.8 ± 6.9 | 13 ± 6.3 |
| Right ventricle minimum pressure, mmHg | 8.9 ± 5.5 | 8 ± 4.9 |
| Mean pulmonary capillary wedge pressure, mmHg | 18.6 ± 7.2 | 19.2 ± 8.6 |
| Maximum pulmonary capillary wedge pressure, mmHg | 23 ± 9.4 | 24.1 ± 12 |
| Left ventricle end systolic pressure, mmHg | 123.5 ± 29.5 | 120.1 ± 21.9 |
| Left ventricle end diastolic pressure, mmHg | 25.1 ± 6.7 | 21.2 ± 7.9 |
| Left ventricle minimum pressure, mmHg | 15.2 ± 6.4 | 12.3 ± 5.7 |
| **Heart imaging measurements** | | |
| Ejection fraction, % | 63.4 ± 8.2 | 31.5 ± 16.1 |
| Left ventricle end diastolic volume, mL | 164.3 ± 59.8 | 281.3 ± 107.3 |
| Left ventricle end systolic volume, mL | 69.5 ± 39.3 | 206.4 ± 105.3 |
| Right ventricle end diastolic volume, mL | 182 ± 77.4 | 195.8 ± 76.4 |
| Right ventricle end systolic volume, mL | 95.3 ± 61.2 | 124.8 ± 69.2 |
| Left ventricle mass, g | 129.8 ± 53.6 | 152 ± 48.3 |
| Right ventricle mass, g | 69 ± 33.8 | 66.7 ± 28.8 |
| Maximum left atrial volume, mL | 81.2 ± 29.4 | 93.7 ± 49.4 |
| Left atrial diameter, end diastolic, cm | 4.4 ± 0.8 | 24.1 ± 0.8 |
| Left ventricle posterior wall thickness, cm | 1.1 ± 0.3 | 1 ± 0.2 |
| Interventricular Septum wall thickness, cm | 1.2 ± 0.4 | 1 ± 0.2 |
| E/A ratio | 1.4 ± 0.7 | 1.6 ± 0.9 |
| Aortic valve peak pressure gradient, mmHg | 14.8 ± 17.0 | 7.4 ± 6.2 |
| Pulmonary valve peak pressure gradient, mmHg | 5.6 ± 5.0 | 3.7 ± 2.2 |
| Mitral valve mean pressure gradient, mmHg | 4.6 ± 2.3 | 3.7 ± 2.0 |
| Mitral valve regurgitation worse than mild | 16(12.5) | 69(32.1) |
| Aortic valve regurgitation worse than mild | 5(3.9) | 10(4.7) |
| Tricuspid valve regurgitation worse than mild | 31(24.2) | 44(20.5) |
| Pulmonary valve regurgitation worse than mild | 7(5.5) | 8(3.7) |

Data presented as mean ± standard deviation for continuous variables, or *n* for categorical variables.

**Supplementary Table 3**
#### Physiological functional parameters and additional simulation outputs

| Physiological functional parameters | | Simulation outputs |
|---|---|---|
| $kact_{LV}$, – | LV active contractility | LVpower, W |
| $kact_{RV}$, – | RV active contractility | LVpowerindex, W/cm$^2$ |
| $kpas_{LV}$, – | LV passive stiffness | RVpower, W |
| $kpas_{RV}$, – | RV passive stiffness | RVpowerindex, W/cm$^2$ |
| $Vw_{LV}$, cm$^3$ | LV free wall volume | LVpower/mass ratio, W/g |
| $Vw_{RV}$, cm$^3$ | RV free wall volume | RVpower/mass ratio, W/g |
| $Vw_{SEP}$, cm$^3$ | Septum free wall volume | $\text{Strain}_{\text{LV}}$, % |
| $Amref_{LV}$, cm$^2$ | LV midwall surface area | $\text{Strain}_{\text{RV}}$, % |
| $Amref_{RV}$, cm$^2$ | RV midwall surface area | $\text{Strain}_{\text{SEP}}$, % |
| $Amref_{SEP}$, cm$^2$ | Septum midwall surface area | $\text{Stress}_{\text{LV}}$, mmHg |
| $\underline{kact_{At}}$, – | Atrial active contractility | $\text{Stress}_{\text{RV}}$, mmHg |
| $\underline{kpas_{At}}$, – | Atrial passive stiffness | $\text{Stress}_{\text{SEP}}$, mmHg |
| $\underline{Pc}$, mmHg | Atrial pressure contributed from collagen | – |
| $\underline{R_{to}}$, mmHg·s/ml | Tricuspid valve resistance open state | – |
| $R_{ao}$, mmHg·s/ml | Aortic valve resistance open state | – |
| $R_{mo}$, mmHg·s/ml | Mitral valve resistance open state | – |
| $R_{po}$, mmHg·s/ml | Pulmonary valve resistance open state | – |
| $R_{tc}$, mmHg·s/ml | Tricuspid valve resistance close state | – |
| $R_{ac}$, mmHg·s/ml | Aortic valve resistance close state | – |
| $R_{mc}$, mmHg·s/ml | Mitral valve resistance close state | – |
| $\underline{R_{pc}}$, mmHg·s/ml | Pulmonary valve resistance close state | – |
| $C_{SV}$, ml/mmHg | Compliance of systemic veins | – |
| $C_{PV}$, ml/mmHg | Compliance of pulmonary veins | – |
| $C_{SA}$, ml/mmHg | Compliance of systemic arteries | – |
| $C_{PA}$, ml/mmHg | Compliance of pulmonary arteries | – |
| $R_{Veins}$, mmHg·s/ml | Resistance of veins | – |
| $R_{SA}$, mmHg·s/ml | Resistance of systemic arteries | – |
| $R_{PA}$, mmHg·s/ml | Resistance of pulmonary arteries | – |
| $R_{tSA}$, mmHg·s/ml | Transmural resistance of systemic arteries | – |
| $R_{tPA}$, mmHg·s/ml | Transmural resistance of pulmonary arteries | – |
| $K_{PC}$, lnmmHg | Exponential pericardium constraint | – |
| $B_{PC}$, mmHg | Constant pericardium constraint | – |

Underlined values represent global constants, distinguishing them from patient-specific parameters.
Additional simulation outputs pertain to variables not included in Supplementary Table 2.

#### 2. Method S1: Cohort Discovery

A retrospective cohort of 343 patients, selected from all patients with HF who visited the University of Michigan Health System (UMHS) between June 1, 2009, and November 30, 2023, was identified using Electronic Health Records (EHR) data. In brief, all adult HF patients without a single ventricle defect and with at least one recorded right heart catheterization (RHC), transthoracic echocardiography (TTE), and cardiac magnetic resonance imaging (CMR) were pre-screened (N=2727). Qualified examinations were defined based on criteria from patients' EHR: RHC must report cardiac output (CO), heart rate (HR), pulmonary artery diastolic pressure (PADP), pulmonary artery systolic pressure (PASP), and pulmonary capillary wedge pressure (PCWP); TTE must report interventricular septum thickness (IVSd), left ventricular posterior wall thickness (LVPWd), left ventricular ejection fraction (LVEF), left ventricular internal diameter at end-diastole (LVIDd), left ventricular internal diameter at end-systole (LVIDs), left atrial diameter (LAd), and E/A ratio; CMR must report left ventricular end-diastolic volume (LVEDV), left ventricular end-systolic volume (LVESV), right ventricular end-diastolic volume (RVEDV), and right ventricular end-systolic volume (RVESV). A 3-month window was defined, within which cardiovascular function was assumed to remain stable. From the pre-screened population, 371 patients with qualified RHC, TTE, and CMR examinations within the specified 3-month window were included in the final cohort (Supplementary Figure 8). Further exclusions were made for patients who were not diagnosed with HF before the time window (N=11); patients who underwent heart transplant or LVAD (left ventricular assist device) implantation within the specified 3-month window (N=3), patients with a history of heart transplant prior to the 3-month window with no evidence of new onset HF (N=7), and patients with a significant shunt, indicated by an abnormal Qp/Qs ratio, defined as greater than 1.2 or less than 0.8, was identified in their RHC/CMR reports (N=11). Finally, clinical variables, including demographics, pre-existing medical conditions, medication history, hemodynamics measurements, heart imaging measurements, laboratory tests, and outcome data (endpoints defined as LVAD implantation, heart transplant, or all-cause mortality), were collected from EHR and double-blind verified by our clinical team. Survival data longer than 5 years are right censored at year 5 (Supplementary Figure 9).

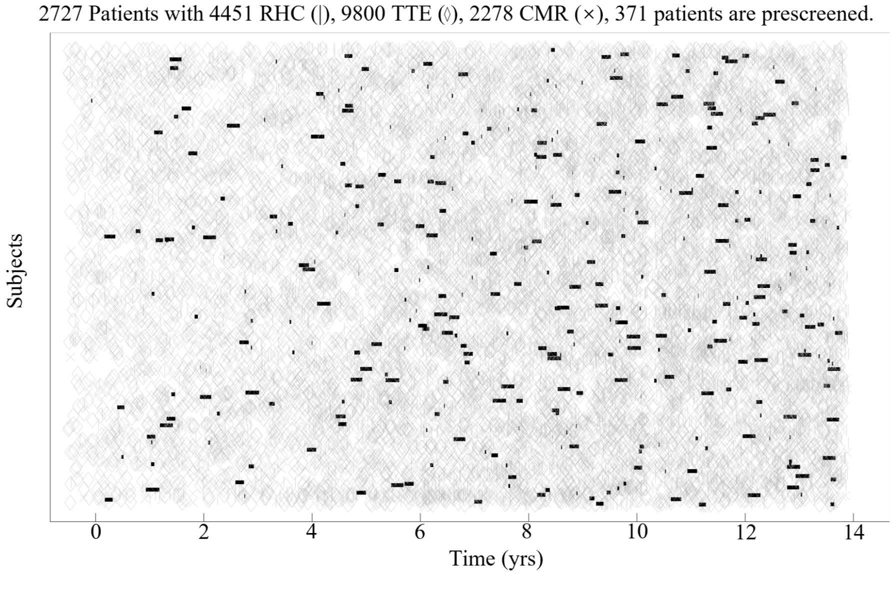


***Supplementary Figure* 8: Heart failure patients identified based on procedure availability within a 3-month window**
The X-axis represents the date, while each row on the Y-axis corresponds to an individual patient. Different symbols represent different types of procedures. A thick black line indicates the time window during which all three procedures occurred. Abbreviations are defined in Table 1.

An independent validation cohort of 113 patients was obtained from the University of Wisconsin (UW)-Madison ('external testing set'). These participants presented to the UW-Madison Pulmonary Hypertension Clinic with worsening dyspnea (New York Heart Association Class II-III) and underwent clinically indicated invasive

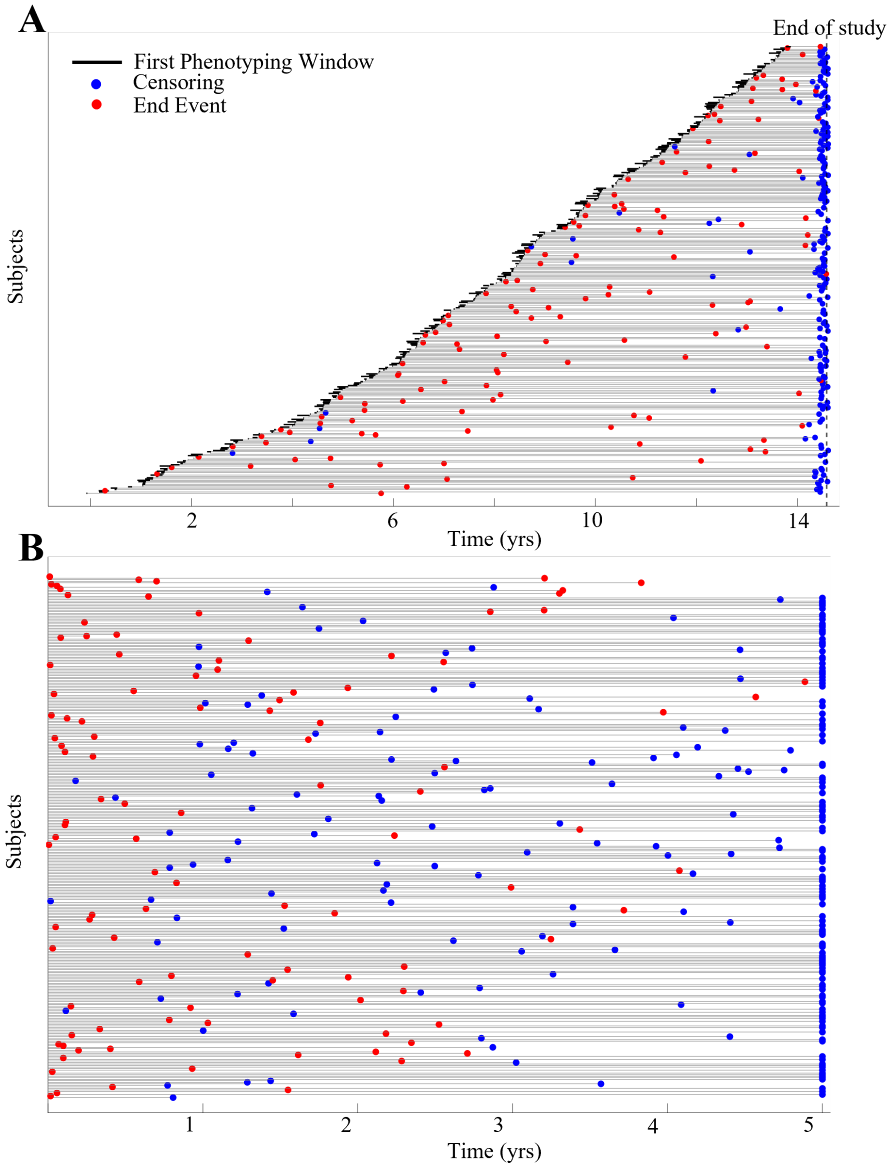


***Supplementary Figure* 9: Overview of outcome data for UMHS patients**
(A) Raw outcome data showing the primary composite endpoint. (B) Data censored at 5 years.

cardiopulmonary exercise testing. Each participant received a comprehensive resting transthoracic echocardiographic evaluation, a six-minute walk test, and a clinical assessment by a PH specialist. Of these, 86 patients had sufficient data (RHC,TTE, and CMR measurements) to identify digital twins. Following the 2022 ERS/ESC guidelines, 32 participants were further diagnosed with HFpEF, defined as having a resting PCWP $\geq$ 15 mmHg and/or an exercise PCWP $\geq$ 25 mmHg (for patients with EF $\geq$ 50%, HF symptoms, and structural, functional, or biomarker evidence of cardiac abnormalities). The follow-up period was one year, with the composite endpoint defined as all-cause mortality and rehospitalization. This study was approved by the UW-Madison institutional review board and adhered to the guidelines of the Declaration of Helsinki.

### 3. Method S2: Digital Twin Identification

#### 3.1. Computational Model

##### *3.1.1. Ventricle Model*

Digital twins are identified using a computational mechanistic model, as described below. This study makes use of the TriSeg cardiac model [1] which represents the heart as three spherical shell sectors joined together, representing the right ventricular free-wall (RW), interventricular septal wall (SW), and left ventricular free-wall (LW). The TriSeg heart geometry is fully described by a wall volume parameter for each segment and four distance parameters describing the position of the three "midwalls". For each of the three spherical segments, the midwall is defined as the imaginary spherical cap surface within the wall that divides the shell into two sub-shells of equal volume (note this is not equal to the surface halfway between the inner and outer surfaces of the wall). To make a valid representation of the heart, the right ventricular midwall, septal midwall, and left ventricular midwall intersect at a junction circle, where they are mechanically coupled and form the enclosed left and right ventricles of the heart. The centers of the midwall surface are collinear, making the TriSeg heart cylindrically symmetric and motivating the creation of the two-dimensional cylindrical coordinate system defined in [1] which four distance parameters $x_{m,LV}$, $x_{m,RV}$, $x_{m,SEP}$, and $y_m$ describe. The TriSeg heart is rotationally symmetric through the axial $x$-axis, which is defined as positive towards the right ventricle, and has a radial $y$ axis. The junction plane is set at $x = 0$, with the three midwall surfaces' boundaries on the same junction circle with center $(0, 0)$ and radius $y_m$. The three $x_{m,i}$ parameters then describe how dilated the midwall surfaces are by measuring the length from the origin to the midwall surface along the $x$-axis (e.g. if $x_{m,i} = y_m$, the midwall surface is a hemisphere).

For any patient, an initial estimate of the seven parameters for wall volume and midwall geometry can be inferred given values for ventricular lumen volume and ventricular wall thickness or mass. The TriSeg heart is then parameterized to match these clinical measurements, which either come from TTE or CMR (Supplementary table 2). Note that the following calculations are to establish initial conditions, and don't necessarily reflect the steady-state model-simulated values of the TriSeg cardiac geometry. To solve for the initial TriSeg geometry, we first examine the left heart, which is the structure formed from the left-ventricular (LV) free wall and the interventricular septum. We make the initial approximation that the union of the LV and septal midwall surfaces forms a sphere, and with a shared lumen volume and wall thicknesses the the LV and septum, the left heart takes the shape of a spherical shell (which may potentially have two different thicknesses). This is expressed below, where $r_{m,i}$ is the radius of the midwall sphere.

$$r_{m,LV} = r_{m,SEP} = r_m$$

We also assume that the septal midwall surface area is one half of the LV midwall surface area. This is expressed below, where $A_{m,ref,i}$ is the area of the midwall surface at end-diastole, which is also used as a parameter for the mechanical model.

$$A_{m,ref,LV} = 2 * A_{m,ref,SEP}$$

Now, with these assumptions, a left-ventricular lumen volume, and either two wall thicknesses or one left heart mass, we can determine the parameters $V_{w,LV}$, $V_{w,SEP}$, $x_{m,LV}$, $x_{m,SEP}$, and $y_m$. Given septal wall thickness $H_{SW}$ and LV wall thickness $H_{LW}$, we obtain the following equations satisfying 1 – the enclosed volume of the left heart is $LVEDV$ – and 2 – the LV and septal midwall surfaces are on the same sphere, where $r_{i,i}$ is the radius from the center of the midwall sphere to the model endocardium.

$$LVEDV = \frac{2\pi}{3}\left(r_{i,SEP}^3(1-\cos(\beta)) + r_{i,LV}^3(1-\cos(\pi-\beta))\right) \tag{1}$$

$$r_{i,SEP}^3 + (r_{i,SEP} + H_{SW})^3 = r_{i,LV}^3 + (r_{i,LV} + H_{LW})^3 \tag{2}$$

With $r_{i,LV}$ and $r_{i,SEP}$, we can determine $r_m$, the radius of the midwall sphere shared by the LV and septum, and $V_{w,LV}$, and $V_{w,SEP}$

$$r_m = \frac{1}{2}((r_{i,LV} + H_{LW})^3 + r_{i,LV}^3)$$

$$V_{w,LV} = \frac{2\pi}{3}(1 - \cos(\pi - \beta))((r_{i,LV} + H_{LW})^3 - r_{i,LV}^3)$$

$$V_{w,SEP} = \frac{2\pi}{3}(1 - \cos(\beta))((r_{i,SEP} + H_{SW})^3 - r_{i,SEP}^3)$$

$\beta$ is the angle swept from $(x_{m,SEP}, 0)$ to $(0, y_m)$ along the left heart midwall sphere:

$$\beta = \cos^{-1}(2 * \frac{A_{m,ref,LV}}{A_{m,ref,LV} + A_{m,ref,SEP}} - 1) = \cos^{-1}(2 * \frac{2}{3} - 1) = \cos^{-1}(\frac{1}{3})$$

If a single left heart mass $LV_m$ (encompassing the LV free wall and interventricular septum) is provided in place of separate wall thicknesses, the same parameters can be calculated more readily. As previously, we assume that the septal midwall surface area is half of that of the left ventricle and further assume a uniform thickness throughout the entire left heart. $V_{w,LV}$ and $V_{w,SEP}$ are readily calculated below:

$$V_{w,LV} = \frac{A_{m,ref,LV}}{A_{m,ref,LV} + A_{m,ref,SEP}} \frac{LV_m}{\rho_{myo}} = \frac{2}{3}\frac{LV_m}{\rho_{myo}}$$

$$V_{w,SEP} = \frac{LV_m}{\rho_{myo}} - V_{w,LV}$$

where $\rho_{myo}$ is the density of the myocardium, taken to be 1.055 g/mL. $r_m$ is then calculated from wall thickness and $LVEDV$:

$$H = \sqrt[3]{r_i^3 + \frac{3}{4\pi}\frac{LV_m}{\rho_{myo}}} - r_i$$

$$r_m = \sqrt[3]{\frac{1}{2}\left(\left(\sqrt[3]{\frac{3}{4\pi}LVEDV} + H\right)^3 + \frac{3}{4\pi}LVEDV\right)}$$

From either set of information, the shared $r_m$ can be used to calculate $y_m$, $x_{m,LV}$, and $x_{m,SEP}$:

$$y_m = r_m \sin\beta \tag{3}$$

$$x_{m,SEP} = r_m - \sqrt{r_m^2 - y_m^2} \tag{4}$$

$$x_{m,LV} = x_{m,SEP} - 2r_m \tag{5}$$

With $V_{w,LV}$, $V_{w,SEP}$, $x_{m,LV}$, $x_{m,SEP}$, and $y_m$, the left heart is fully described. Adding right-ventricular (RV) free wall volume $V_{w,RV}$ and the RV midwall distance $x_{m,RV}$, the full cardiac geometry is captured. We infer these parameters with RV lumen volume $RVEDV$ and thickness $H_{RW}$ (end-diastole) or free wall mass $RV_m$. The right heart geometry should be parameterized such that the RV midwall surface intersects the junction plane at $(0, \pm y_m)$ and that RV lumen has volume $RVEDV$ while having a portion of the septum protruding into the positive $x_m$ domain. This latter constraint is expressed below:

$$RVEDV = V_{m,RV} - \frac{1}{2}V_{w,RV} - V_{sep} \tag{6}$$

where $V_{m,RV}$ is the volume of the spherical cap enclosed between the midwall surface and junction plane, and $V_{sep}$ is the volume of the septal spherical cap that protrudes into the right side of the TriSeg heart (already known from the left heart parameterization):

$$V_{sep} = V_{m,SEP} + \frac{1}{2}V_{w,SEP}$$

We now introduce the auxiliary variable $h$, defined as the distance along the $x$ axis (towards the left heart) from the origin to the RV midwall sphere, if the surface was extended beyond the junction plane to form a complete sphere. Note that since $h$ is in the negative $x$ domain, $h$ is defined to be less than 0. Given RV wall mass or thickness, $RVEDV$, and a description of the left heart, fixing a value of $h$ fully captures the right heart geometry. To solve for $h$, we express $V_{w,RV}$ and $V_{m,RV}$ in terms of $h$, then substitute these expressions into 6.

First, $x_{m,RV}$ and $r_{m,RV}$ are given by

$$\begin{aligned} x_{m,RV} &= -\frac{y_m^2}{h} \\ r_{m,RV} &= -\frac{(h^2 + y_m^2)}{2h} \end{aligned} \tag{7}$$

With $x_{m,RV}$, we are able to calculate $V_{m,RV}$ using spherical cap equations (also equation 9 in [1])

$$V_{m,RV} = \frac{\pi}{6}x_{m,RV}(x_{m,RV}^2 + 3y_m^2) = -\frac{\pi}{6}\frac{y_m^2}{h}\left(\left(\frac{y_m^2}{h}\right)^2 + 3y_m^2\right) \tag{8}$$

With midwall volume $V_{m,RV}$ in terms of $h$, wall volume $V_{w,RV}$ can be calculated using integration in spherical coordinates, choosing bounds expressed in terms of $h$.

$$\begin{aligned} V_{w,RV} &= \frac{2\pi}{3}(1 + \frac{h^2 - y_m^2}{h^2 + y_m^2})((r_{m,RV} + \delta_{RW})^3 - (r_{m,RV} + \delta_{RW} - H_{RW})^3) \\ \delta_{RW} &= \frac{1}{2}\left(\sqrt[3]{\sqrt{H_{RW}^6 + 16r_{m,RV}^6} + 4r_{m,RV}^3} - \frac{H_{RW}^2}{\sqrt[3]{\sqrt{H_{RW}^6 + 16r_{m,RV}^6} + 4r_{m,RV}^3}} + H_{RW} - 2r_{m,RV}\right) \end{aligned} \tag{9}$$

Here $\delta_{RW}$ is the distance from the RV midwall sphere to the model epicardium (note that this is not half of $H_{RW}$). Now that $V_{w,RV}$ is in terms of $h$ (since $r_{m,RV}$ and $\delta_{RW}$ are both in terms of $h$), 6 can be numerically solved for $h$.

If instead of wall thickness $H_{RW}$ we start with RV wall mass $RV_m$, we again solve for $h$ using 6, with a much simpler expression for $V_{w,RV}$

$$V_{w,RV} = \frac{RV_m}{\rho_{myo}}$$

With $h$ obtained from either set of input data, the values for $x_{m,RV}$ and $V_{w,RV}$ (if given $H_{RW}$) can finally be determined from 7 and 9, thus completing the parameterization of the TriSeg heart.

This geometric representation of the heart is coupled to a mechanical model that governs stress, tension, and ultimately chamber pressure development through the cardiac cycle. Our model adopts simplifications of [1]'s formulation of passive myofiber stress development and cardiac activation that were previously used in [2].

Myofiber stress ($\sigma$) in each wall segment is modeled as a linear combination of passive and active stress as in [1].

$$\sigma = k_{pas}\sigma_{pas} + k_{act}\sigma_{act} \tag{10}$$

$k_{pas}$ and $k_{act}$ are the unitless constants that scale the passive and active stresses. Passive ($\sigma_{pas}$) and active ($\sigma_{pas}$) stress develop according to cardiac activation $Y(t)$ and sarcomere length. At any given time, the total sarcomere length in a wall segment is calculated from the geometry of that wall segment, and subsequently used to obtain sarcomere strain, then passive stress. The sarcomere also has an active contractile element that changes length according to a first-order ODE, which is combined with the sarcomere strain and cardiac activation to obtain active stress.

First, the myofiber strain $\epsilon$ is calculated below (from equation 15 of [1])

$$\epsilon \approx 0.5 \ln\left(\frac{A_m}{A_{m,ref}}\right) - \frac{1}{12} z^2 - 0.019 z^4 \qquad \text{where} \qquad z = \frac{3 C_m V_w}{2 A_m} \tag{11}$$

$A_m$ is midwall surface area, $C_m$ is the midwall curvature (reciprocal of the radius $r_m$), and $z$ is a dimensionless curvature parameter related to the ratio of wall thickness to midwall radius (each of these are calculated from the distance parameters $x_{m,i}$ and $y_m$). Sarcomere length is then calculated as

$$L_s = L_{s,ref} e^{\epsilon} \tag{12}$$

$L_{s,ref}$ is the reference sarcomere length at end-diastole, 2.0 $\mu$m. As in [2], passive stress is formulated based on "the exponential formula from [3]":

$$\sigma_{pas} = k_{pas} \left(\frac{L_s}{L_{sc,0}} - 1\right)^{\gamma} \tag{13}$$

$L_{sc,0}$ is the length of the contractile element of the sarcomere model at zero stress (TABLE) and $\gamma$ is a parameter that dictates stress development.

Active stress $\sigma_{act}$ depends on both the sarcomere length $L_s$ from the model geometry and the contractile element length $L_{sc}$, which varies with time according to

$$\dot{L_{sc}} = \left(\frac{L_s - L_{sc}}{L_{se,iso}} - 1\right) v_{max} \tag{14}$$

$L_{se,iso}$ is the length of the series elastic element of the sarcomere when isometrically stress (zero strain), and $v_{max}$ is the contractile element shortening velocity at zero load (TABLE). $\sigma_{act}$ also depends on a mechanical activation function $Y(t)$ that drives the cardiac cycle in the ventricles. As in [2], we model the activation function as a piecewise sinusoidal function:

$$Y(t) = \begin{cases} 0.5\left(1 - \cos\left(\frac{\pi t}{k_{TS}}\right)\right) & 0 \leq t \leq k_{TS} \\ 0.5\left(1 + \cos\left(\frac{\pi (t - k_{TS})}{k_{TR}}\right)\right) & k_{TS} \leq t \leq k_{TS} + k_{TR} \\ 0 & \text{otherwise} \end{cases} \tag{15}$$

$k_{TS}$ and $k_{TR}$ represent the proportions of the cardiac cycle spent in systole and diastole, respectively. We set $k_{TS}$ to 0.35, and $k_{TR}$ to 0.15 [4]. Finally, the active stress $\sigma_{act}$ is given by:

$$\sigma_{act} = k_{act} Y(t) \frac{L_{sc} - L_{sc,0}}{L_{sc,0}} \frac{L_s - L_{sc}}{L_{sc,0}} \tag{16}$$

Note that these equations are used for each of the three segments of the TriSeg heart. All parameters for cardiac mechanics (excluding the wall segment geometry parameters) are constant across the three segments, except for the scaling parameters $k_{pas}$ and $k_{act}$. These parameters are inferred prior to simulation from end-diastolic and end-systolic chamber pressures, respectively, and therefore have different values for the right and left heart.

$$k_{pas} = \frac{EDP}{\Gamma_d \sigma_{pas,d}}$$
$$k_{act} = \frac{ESP}{\Gamma_s \sigma_{act,s}}$$
$$\Gamma = -\frac{2}{3}\left(1 + \frac{1}{3} z_i^2 + \frac{1}{5} z_i^4\right)$$

Here, $\Gamma$ is a helpful auxiliary variable obtained by backcalculating for chamber pressure from [1] equation 14, using [1]'s equations 12 and 16 (as derived in the supplement of [2]). For both ventricles, $EDP$ and $ESP$ are clinical measures or estimates of ventricular chamber pressures. $\sigma_{pas,d}$ is the passive myofiber stress at end-diastole, and is estimated assuming that $\sigma_{act,d}$ is 0 and that sarcomere length at end-diastole is $2\mu$m. Similarly, $\sigma_{act,s}$ is the active myofiber stress at end-systole and is estimated assuming 60% activation ($Y(t) = 0.6$) at end-systole, and that $\sigma_{pas,s}$ is 0. The $k$ parameters are shared between the LV free wall and septal segments: $k_{pas,LV}$ and $k_{act,LV}$ are calculated based on LV free wall geometry, but used for both the LV free wall and septum in calculating the total myofiber stress $\sigma$ during simulation. <Note that $k_{pas,LV}$, $k_{pas,RV}$, $k_{act,LV}$, and $k_{act,LV}$ are optimized for each patient.>

Now with myofiber stress and wall geometry, we obtain midwall tension $T_m$ as in [1] equation 16:

$$T_m = \frac{V_w \sigma}{2A_m}\left(1 + \frac{z^2}{3} + \frac{z^4}{5}\right) \tag{17}$$

Midwall tension can then be resolved into axial tension $T_x$ and radial tension $T_y$. Finally, chamber pressures can be calculated using the law of Laplace (equation 14 of [1]):

$$P_V = \frac{2T_x}{y_m} + P_{\text{peri}} \tag{18}$$

The mechanical coupling of the TriSeg heart occurs at the junction circle (see 3.1.1); $T_x$ and $T_y$ across the three segments sums to 0 force.

$$T_{x,LW} + T_{x,SW} + T_{x,RW} = 0 \tag{19}$$

$$T_{y,LW} + T_{y,SW} + T_{y,RW} = 0 \tag{20}$$

Additionally, the lumen volumes of the LV and RV are also balanced:

$$-V_{LV} - 0.5V_{w,LW} - 0.5V_{w,SW} + V_{m,SW} - V_{w,LW} = 0 \tag{21}$$

$$V_{RV} + 0.5V_{w,RW} + 0.5V_{w,SW} + V_{m,SW} - V_{w,RW} = 0 \tag{22}$$

##### *3.1.2. Atrial and Pericardial Model*

Building on the TriSeg model as implemented in [1] and [2], we introduced simple models of the atria to more accurately simulate transvalvular blood flow, enhancing the representation of cardiac hemodynamics. The atria are modeled as time-varying elastic compartments, with an activation function $Y_a(t)$offset from the ventricular activation function by 8% of the cardiac cycle, indicating that the atria contract prior to the ventricles.

$$P_a = E_{pas}(V_a - V_{0,a}) + E_{act}(V_a - V_{0,a})Y_a(t) + P_c \exp\left(\frac{V_a - V_{0c,a}}{V_{1c,a}}\right) + P_{peri} \tag{23}$$

Here, we express the atrial pressure, like the ventricular pressure, as a weighted sum of active and passive pressure development, in this case including contributions from elastin, collagen, and the pericardium. The $E$ parameters scale the elastance of the tissue, as does the $P_c$ term which scales the exponential collagen-based pressure. $V_a$ represents the current atrial volume, $V_{0,a}$ represents an atrial zero-pressure volume, and $V_{0c,a}$ and $V_{1c,a}$ dictate the volume threshold and rate of the exponential collagen-based pressure increase, respectively. Finally, the time-dependence comes from the atrial activation function $Y_a(t)$, which is a gaussian function shared across all simulations with a maximum that is shifted slightly earlier from peak ventricular activation. This model is identical for the left and right atria, except for the $V_{0,a}$ and $V_{0c,a}$ are derived from each patient's estimated (or measured) minimal and maximal atrial volume, respectively.

We also introduced a simple pericardial model, different from that used in [2], to better fit the RV EDPVR:

$$P_{peri} = \exp\left(\frac{V_{total}}{V_{0,total}} K_{PC}\right) + B_{PC} \tag{24}$$

Pericardial pressure exponentially increases according to $K_{PC}$ as the current total heart volume $V_{total}$ exceeds $V_{0,total}$. $V_{0,total}$ is calculated for each individual as the maximal total heart volume: the sum of ventricular and atrial chamber volumes in end-diastole and wall volumes. The last term, $B_{PC}$ - constant pericardium constraint - acts as a thoracic baseline pressure. Both $B_{PC}$ and $K_{PC}$ are among the set of optimized parameters, and are adjusted during the optimization process for each patient.

##### *3.1.3. Valvular and Vascular Model*

The heart model outlined above – the ventricles, atria, and pericardium – are integrated with a lumped-parameter vascular model and valvular model to make a complete, closed-loop cardiovascular model. The vascular model is divided into four compartments: systemic and pulmonary arteries (SA and PA) and systemic and pulmonary veins (SV and PV). Each compartment is characterized by its compliance and resistance to both inflow and outflow, with the SA and PA also having additional transmural resistance, as shown in Figure 2A.

We calculate compartmental compliances ($C_j$, mL mmHg-1) and intercompartmental resistances ($R_j$, mmHg s mL-1) based on change in volumes and pressures. That is, $C_j$ is the ratio of the compartmental stressed volume ($V_{s,j}$) to the compartmental pressure ($P_j$ mmHg):

$$C_j = \frac{V_{s,j}}{P_j} \tag{25}$$

$R_j$ is the ratio of the pressure drop across the resistance to the corresponding flow ($Q_j$, mL s-1):

$$R_j = \frac{P_j - P_{j-1}}{Q_j} \tag{26}$$

As many patient pathologies included valvular disease, we adjusted the model to be able to simulate valvular regurgitation and stenosis. This was accomplished by making the valve components into resistors instead of diodes, where instead of switching between completely open ($R_o$) and completely closed ($R_c$), each valve has a forward and backward resistance to flow, where a higher forward resistance simulates stenosis and a lower backward resistance simulates regurgitation or backflow. The valve resistances are similarly computed as a ratio of pressure difference and flow across the valve with bi-direction.

To solve the above compliance and resistance, the circulation model is formulated such that the circulating volume is the sum of the stressed volume for all compartments. Volume is conserved by formulating differential equations using Kirchhoff's law as:

$$\frac{dV_{s,j}}{dt} = Q_{in} - Q_{out} \tag{27}$$

where $Q_{in}$ and $Q_{out}$ are the time-dependent flows in and out of each compartment.

For the atrial compartments, the inlet flow are described as:

$$Q_{in,LA} = \frac{P_{PV} - P_{LA}}{R_{Veins}} \tag{28}$$

$$Q_{in,RA} = \frac{P_{SV} - P_{RA}}{R_{Veins}} \tag{29}$$

where we assume all venous compartments have the same resistance.

For the ventricle compartments, the inlet flow are described as:

$$Q_{in,LV} = \begin{cases} \frac{P_{LA}-P_{LV}}{R_{mo}} & P_{LA} \geq P_{LV} \\ \frac{P_{LA}-P_{LV}}{R_{mc}} & P_{LA} < P_{LV} \end{cases} \tag{30}$$

$$Q_{in,RV} = \begin{cases} \frac{P_{RA}-P_{RV}}{R_{to}} & P_{RA} \geq P_{RV} \\ \frac{P_{RA}-P_{RV}}{R_{tc}} & P_{RA} < P_{RV} \end{cases} \tag{31}$$

where The subscripts $t$ and $m$ represent the mitral valve and tricuspid valve, respectively. And the outlet flow can be described as:

$$Q_{out,LV} = \begin{cases} \frac{-R_{SA}V_{SA}+C_{SA}P_{LV}R_{SA}+C_{SA}P_{LV}R_{tSA}-C_{SA}P_{SV}R_{tSA}}{C_{SA}(R_{SA}R_{ao}+R_{SA}R_{tSA}+R_{ao}R_{tSA})} & P_{LV} \geq P_{SA} \\ \frac{-R_{SA}V_{SA}+C_{SA}P_{LV}R_{SA}+C_{SA}P_{LV}R_{tSA}-C_{SA}P_{SV}R_{tSA}}{C_{SA}(R_{SA}R_{ac}+R_{SA}R_{tSA}+R_{ac}R_{tSA})} & P_{LV} < P_{SA} \text{ with aortic valve regurgitation} \\ 0 & P_{LV} < P_{SA} \text{ without aortic valve regurgitation} \end{cases} \tag{32}$$

$$Q_{out,RV} = \begin{cases} \frac{-R_{PA}V_{PA}+C_{PA}P_{RV}R_{PA}+C_{PA}P_{RV}R_{tPA}-C_{PA}P_{PV}R_{tPA}}{C_{PA}(R_{PA}R_{po}+R_{PA}R_{tPA}+R_{po}R_{tPA})} & P_{RV} \geq P_{PA} \\ \frac{-R_{PA}V_{PA}+C_{PA}P_{RV}R_{PA}+C_{PA}P_{RV}R_{tPA}-C_{PA}P_{PV}R_{tPA}}{C_{PA}(R_{PA}R_{pc}+R_{PA}R_{tPA}+R_{pc}R_{tPA})} & P_{RV} < P_{PA} \text{ with pulmonary valve regurgitation} \\ 0 & P_{RV} < P_{PA} \text{ without pulmonary valve regurgitation} \end{cases} \tag{33}$$

The corresponding arterial compartments may also be represented as follows:

$$P_{SA} = \begin{cases} \frac{R_{SA}R_{ao}V_{SA}+C_{SA}P_{LV}R_{SA}R_{tSA}+C_{SA}P_{SV}R_{ao}R_{tSA}}{C_{SA}(R_{SA}R_{ao}+R_{SA}R_{tSA}+R_{ao}R_{tSA})} & P_{LV} \geq P_{SA} \\ \frac{R_{SA}R_{ac}V_{SA}+C_{SA}P_{LV}R_{SA}R_{tSA}+C_{SA}P_{SV}R_{ac}R_{tSA}}{C_{SA}(R_{SA}R_{ac}+R_{SA}R_{tSA}+R_{ac}R_{tSA})} & P_{LV} < P_{SA} \text{ with aortic valve regurgitation} \\ \frac{R_{SA}V_{SA}+C_{SA}P_{SV}R_{tSA}}{C_{SA}(R_{SA}+R_{tSA})} & P_{LV} < P_{SA} \text{ without aortic valve regurgitation} \end{cases} \tag{34}$$

$$Q_{SA} = \begin{cases} \frac{R_{ao}V_{SA}-C_{SA}P_{SV}R_{ao}+C_{SA}P_{LV}R_{tSA}-C_{SA}P_{SV}R_{tSA}}{C_{SA}(R_{SA}R_{ao}+R_{SA}R_{tSA}+R_{ao}R_{tSA})} & P_{LV} \geq P_{SA} \\ \frac{R_{ac}V_{SA}-C_{SA}P_{SV}R_{ac}+C_{SA}P_{LV}R_{tSA}-C_{SA}P_{SV}R_{tSA}}{C_{SA}(R_{SA}R_{ac}+R_{SA}R_{tSA}+R_{ac}R_{tSA})} & P_{LV} < P_{SA} \text{ with aortic valve regurgitation} \\ \frac{V_{SA}-C_{SA}P_{SV}}{C_{SA}(R_{SA}+R_{tSA})} & P_{LV} < P_{SA} \text{ without aortic valve regurgitation} \end{cases} \tag{35}$$

$$P_{PA} = \begin{cases} \frac{R_{PA}R_{po}V_{PA}+C_{PA}P_{RV}R_{PA}R_{tPA}+C_{PA}P_{PV}R_{po}R_{tPA}}{C_{PA}(R_{PA}R_{po}+R_{PA}R_{tPA}+R_{po}R_{tPA})} & P_{RV} \geq P_{PA} \\ \frac{R_{PA}R_{pc}V_{PA}+C_{PA}P_{RV}R_{PA}R_{tPA}+C_{PA}P_{PV}R_{pc}R_{tPA}}{C_{PA}(R_{PA}R_{pc}+R_{PA}R_{tPA}+R_{pc}R_{tPA})} & P_{RV} < P_{PA} \text{ with pulmonary valve regurgitation} \\ \frac{R_{PA}V_{PA}+C_{PA}P_{PV}R_{tPA}}{C_{PA}(R_{PA}+R_{tPA})} & P_{RV} < P_{PA} \text{ without pulmonary valve regurgitation} \end{cases} \tag{36}$$

$$Q_{PA} = \begin{cases} \frac{R_{po}V_{PA}-C_{PA}P_{PV}R_{po}-C_{PA}P_{PV}R_{tPA}+C_{PA}P_{RV}R_{tPA}}{C_{PA}(R_{PA}R_{po}+R_{PA}R_{tPA}+R_{po}R_{tPA})} & P_{RV} \geq P_{PA} \\ \frac{R_{pc}V_{PA}-C_{PA}P_{PV}R_{pc}-C_{PA}P_{PV}R_{tPA}+C_{PA}P_{RV}R_{tPA}}{C_{PA}(R_{PA}R_{pc}+R_{PA}R_{tPA}+R_{pc}R_{tPA})} & P_{RV} < P_{PA} \text{ with pulmonary valve regurgitation} \\ \frac{V_{PA}-C_{PA}P_{PV}}{C_{PA}(R_{PA}+R_{tPA})} & P_{RV} < P_{PA} \text{ without pulmonary valve regurgitation} \end{cases} \tag{37}$$

By computing the derivatives of flow and sarcomere length with the constraint conditions equations 19, 20, 21, and 22, the model can be solved.

#### 3.2. Strategy of Patient-specific Simulation

##### *3.2.1. Parameters Optimization*

Clinical data used as inputs for the computational mechanistic model are summarized in Supplementary Table 2. Hemodynamic measurements were obtained invasively, while geometrical measurements relied on CMR data rather than TTE, with the exception of EF. CMR provides a direct measurement of EF, whereas TTE calculates it based on flow. Therefore, we retained the EF from TTE and used other volumetric data from CMR. For patients with sufficient data—defined as having at least one qualifying RHC, TTE, and CMR report within a specified time window (Figure 1)—the input data was intelligently parsed. This parsed data serves as the target data for the model, guiding the optimization process.

The optimized functional parameters include $V_{w,LV}$, $w_{w,SEP}$, $k_{act}$, $k_{pas}$ for each ventricle, $C$ and $R$ for each vessel compartment, $R_t$ for arteries, $K_{PC}$ and $B_{PC}$ for the pericardium, and $R_{mo}$ (for simulating the E/A ratio). Additional parameters are optimized based on specific patient conditions. For instance, if the patient has valve insufficiency, the corresponding valve's $R_o$ (for stenosis) or $R_c$ (for regurgitation) is optimized. If RV thickness or mass is available, $Vw_{RV}$ is also optimized.

Initial conditions for the geometrical and mechanical parameters are estimated based on section 3.1.1, while $C$ and $R$ are determined using equations 25 and 26. If $R_o$ or $R_c$ require optimization, their initial values are estimated based on the degree of insufficiency to simulate the corresponding cross-valve pressure gradient or regurgitation fraction (RF). If not optimized, $R_{mc}$, $R_{ac}$, and $R_{tc}$ are assigned a value of 65,535—a sufficiently large number to ensure an RF of 0%—while $R_o$ values are set to small numbers close to zero to simulate cross-valve gradients smaller than 2 mmHg. Initial conditions of both $K_{PC}$ and $B_{PC}$ are set to 1, and $R_{Veins}$ is set to 0.04. Similarly, $R_{tSA}$ and $R_{tPA}$ are initialized to 0.08 and 0.01, respectively, based on prior experience. These parameters are then optimized during the simulation.

Moreover, to construct the closed-lumped model described in section 3.1, certain parameters lack sufficient information for precise optimization, so they are assigned global constants. The objective is not to obtain exact representative values but to generate reasonable simulation outputs. Specifically, $E_{pas}$ is set to 0.05, $E_{act}$ to 2.6, $P_c$ to 10, and $V_{1c,a}$ to 5. The other parameters in the atrial model are still not optimized and are instead derived through data imputation. The value of $V_{0c,a}$ is set to the maximum volume of the left atrium, which can either be directly measured or estimated based on LAd [5]. The value of $V_{0,a}$ is set to 0.9 times the minimum volume of the left or right atrium. The minimum volume of the left atrium is based on the assumption that atrial stroke volume accounts for 60% of the ventricular stroke volume. The minimum right atrial volume is determined by a proportional relationship with the left atrial minimum volume. In males, this ratio is 32/25, and in females, it is 23/22, reflecting the ratio of LAmin to RAmin volumes derived from canonical subjects. Total blood volume is calculated based on Nader et al. [6]. After accounting for the volume assigned to the heart model, the remaining blood volume is distributed across the SA, PA, SV, and PV compartments in a ratio of 15%, 5%, 60%, and 20%, respectively. In the veins, 90% of the blood is considered unstressed, while in the SA and PA, 70% and 10%, respectively, are considered unstressed.

This framework ensures that the number of optimized model parameters is always smaller than the number of simulation targets, guaranteeing a unique solution for each patient. Depending on individual data availability, the model used 19 to 37 clinical characteristics to identify 16 to 23 physiological functional parameters. Ultimately, this enables the generation of patient-specific steady-state simulations. Finally, this approach ensured that the framework was able to output 29 physiological functional parameters ($V4c$, $Amref_{LV}$, $Amref_{RV}$, and $Amref_{SEP}$ are not optimized, as they can be directly calculated. Similarly, $C_{PV}$ and $C_{SV}$ are not optimized because, unlike the arteries where changes in volume are reflected by stroke volume, there is insufficient information to accurately determine venous compliance. However, these six parameters remain patient-specific) for the cardiovascular system and produce 50 simulations as their digital twins.

For each patient, The model's functional parameters are optimized by minimizing the weighted squared error between the simulated outputs and the corresponding measured pressures in the ventricles and systemic and pulmonary circulations, as well as the measured volumes and flows in the cardiovascular chambers and vessels. To achieve this, we employ a combination of Genetic Algorithm (*ga*), *patternsearch*, and *fminsearch* optimization functions from MATLAB (MathWorks Inc.), chosen for their effectiveness in navigating the nonlinear, multi-dimensional optimization landscape..

##### 3.2.2. Additional Constraint for the Dicrotic Notch

Although the model includes a valve compartment, it cannot fully replicate the real closing and opening dynamics of the valves, which may manifest as a dicrotic notch in the arterial blood pressure waveform. To address this limitation, we use $R_{tPA}$ and $R_{tSA}$ to simulate this phenomenon. Additionally, two specific targets are incorporated into the loss function during optimization. One target is the dicrotic notch in the systemic arterial blood pressure, calculated using the formula:

$$\mathrm{DNA} = 1.32\left(\frac{\mathrm{SBP} - \mathrm{DBP}}{3} + \mathrm{DBP}\right) - 22.6 \tag{38}$$

where SBP is the systolic blood pressure and DBP is the diastolic blood pressure [7]. The second target is the dicrotic notch in the pulmonary artery, calculated as:

$$\mathrm{DNP} = 1.004\left(\frac{\mathrm{PASP} - \mathrm{PADP}}{3} + \mathrm{PADP}\right) - 0.5 \tag{39}$$

where PASP and PADP represent the pulmonary artery systolic and diastolic pressures, respectively [8]. These formulas are integrated into the optimization process to improve the physiological accuracy of the simulated waveform.

###### *3.2.3. Inconsistency in LV Mass Measurement between TTE and CMR*

During optimization, we identified a scenario where an artificial bias could be introduced, separating patients into two groups based on whether they had CMR data. Specifically, for patients with both LV mass measured by CMR and LV wall thickness measured by TTE, the heart model's geometry incorporates information from both modalities. This can result in LVPWd simulation outputs deviating from target values. In contrast, for patients without CMR, this deviation does not occur (Supplementary Figure 10). The underlying issue is that patients with CMR data use a single simulation to match two targets (LV mass and wall thickness), while patients without CMR data use one target alone.

The aim is not to determine which measurement is more accurate but to minimize this bias while preserving as much measurement information as possible. For patients without CMR, we assume that, if CMR data were available, their LVPWd (Supplementary Figure 10A) and IVSd (data not shown) would follow the same linear relationships observed in those with CMR. Therefore, we calculate assumed LVPWd and IVSd values based on the following relationships:

$$\text{Assumed LVPWd} = 0.4235 \times \text{Real LVPWd} + 0.2021 \tag{40}$$

$$\text{Assumed IVSd} = 0.6342 \times \text{Real IVSd} - 0.003236 \tag{41}$$

These assumed LVPWd and IVSd values are then input into the heart model (3.1.1) to calculate an assumed LV mass.

In summary, for patients with both CMR and TTE measurements, LV mass, LVPWd, and IVSd are included as targets in the loss function during optimization. For patients without CMR, we use the assumed LV mass, along with the real LVPWd and IVSd, as targets in the loss function.

###### *3.2.4. Simulating Cardiovascular System Function in Patients without RV Mass Data*

Most patients in the UMHS cohort lack data on RV free wall volume ($Vw_{RV}$), which could be measured by CMR (as RV mass) or by TTE (as RV thickness). The absence of this measurement impedes building a complete computational model for identifying digital twins. To address this issue, we developed the following workflow.

We employed a lasso regression model to estimate $kpas_{RV}$ and then performed backward imputation of $Vw_{RV}$ based on Section 3.1.1. For training, we selected 34 patients from the UW cohort with RV thickness or mass measurements, while 12 patients from the UMHS cohort with RV mass data were used as the testing set. After obtaining the actual $kpas_{RV}$ values through optimization, we built the lasso regression model to estimate $kpas_{RV}$. The model was trained using five-fold cross-validation, with input features including EF, PASP, PADP, PCWP, CO, SP, BP, LVPWd, and IVSd. The results are presented in Supplementary Figure 11. While the regression performed reasonably well on the training set, it did not generalize effectively to the testing set.

Further analysis showed that the lasso regression model overestimated $kpas_{RV}$ by more than sevenfold in male subjects and elevenfold in female subjects when compared to the values obtained from healthy male and female reference subjects. To correct for this, we used these overestimation factors as calibration coefficients. Specifically,

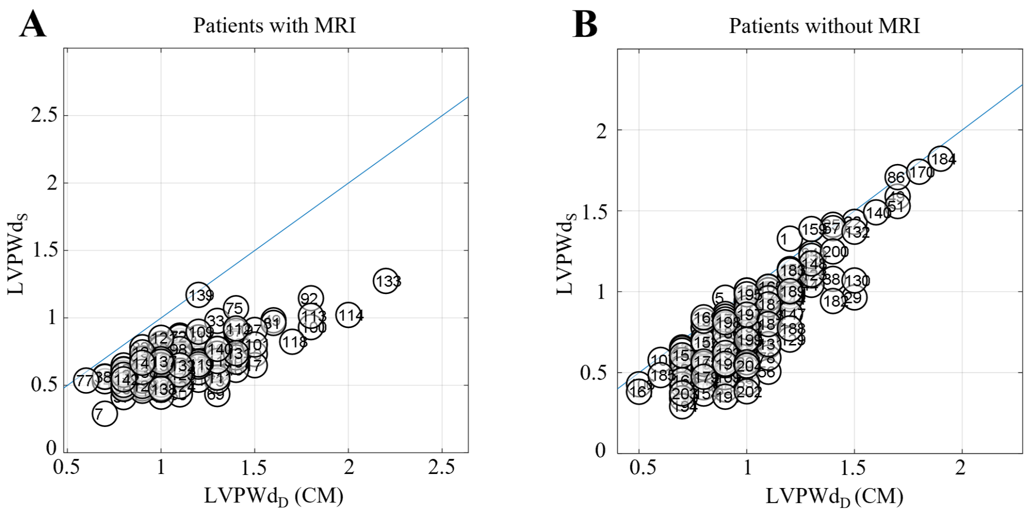


***Supplementary Figure*** **10: Comparison of simulation outputs for LV wall mass and thickness in patients with and without CMR data**

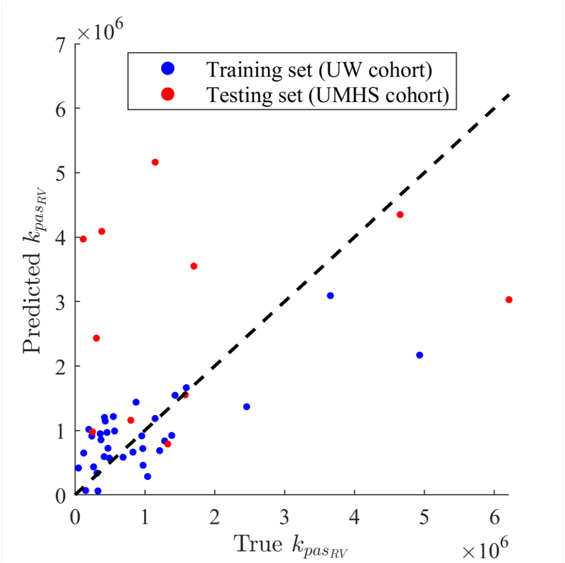


***Supplementary Figure*** **11: Lasso regression-based imputation of $kpas_{RV}$**
Unlike other predictive models, the UW cohort is used as the training set, while the UMHS cohort serves as the external testing set.

after using lasso regression to estimate $kpas_{RV}$ for other patients, we divided the male estimates by the sevenfold factor and the female estimates by the elevenfold factor before performing the backward imputation of $Vw_{RV}$. For patients without direct RV free wall thickness or mass measurements, the estimated $Vw_{RV}$ remains unchanged during optimization.

There are two key points to highlight: First, The primary goal of the above approach is not to achieve precise imputation of $kpas_{RV}$ or $Vw_{RV}$, but rather to provide a physiologically reasonable and consistent assumption for the right ventricle. We initially attempted to use lasso regression to impute RV mass or thickness directly and calibrate using canonical subjects. However, due to differences between the UMHS and UW cohorts—particularly the higher prevalence of right heart complications and hypertrophy in the UW cohort—the regression model often predicted values outside the expected physiological range for the UMHS patients, who generally have less hypertrophic right ventricles. In the UW cohort, RV thickness is typically greater than 1 cm, meaning that the linear relationship derived from the UW data is mostly valid for patients with more severe hypertrophy. In contrast, such cases are much rarer in the UMHS cohort. As a result, many UMHS patients fall outside this range, and their predicted values often lie below the lower bound of the regression line derived from the UW cohort. Although the majority of estimated RV free wall thicknesses fell within reasonable ranges, some extreme values, such as 3 cm thickness, were observed. Therefore, while backward imputation may not yield accurate results, it provides a physiologically plausible assumption. Second, despite this assumption being reasonable, the information derived from the right ventricle remains limited and less reliable. The behavior of a single digital twin model may represent a combination of several unaccounted-for RV phenotypes.

#### 4. Method S3: Clustering Methodology and Interpretation of 3D Volcano Plot

The entire method was developed in the MATLAB environment. Not to develop a more advanced clustering technique, but to identify distinguished and interpretable clusters among patients with HF, we established the following workflow based on functional parameters.

##### 4.1. Data Preprocessing and Determination of Cluster Number

The computational model, based on data from 20-year-old canonical healthy males and females, was first constructed to serve as the reference level (Supplementary Figure 12). Functional parameters related to volume from patients were initially normalized by body surface area (BSA) to account for variations in blood volume, and then all parameters were divided by the corresponding values from a canonical subject. We used all 29 functional parameters as input features for clustering, excluding four $R_c$ parameters. These parameters were excluded because, after normalization, their values ranged from $10^{-10}$ to 1, which we believe could skew the downstream analysis. Instead, we replaced the 4 $R_c$ parameters with regurigation fraction (RF) from the valves, as they likely capture similar characteristics. Thus, we clustered using 25 relative functional parameters along with 4 RF parameters.

Each feature was z-scored, and outliers (128 out of 9947) were defined as any z-score greater than 3 or less than -3. These were replaced by the maximum or minimum values within the non-outlier range. PCA was then performed, not for dimensionality reduction but to project the 29-dimensional data into a 2D space for visualization.

To determine the number of clusters in the UMHS cohort, we first performed hierarchical clustering and examined the dendrogram (Supplementary Figure 13B). Even with functional parameters, HF remains highly heterogeneous, making it difficult to distinguish clear clusters within the cohort. We then used MATLAB's built-in function 'evalclusters' to optimize the cluster number. Both the Davies-Bouldin index and GAP statistic were applied to evaluate clustering performance for $n = 1$ to $n = 25$, using k-means, hierarchical clustering, and Gaussian Mixture Model (GMM) (Supplementary Figure 13A). The DBI measures the ratio of within-cluster dispersion to between-cluster separation, while the gap statistic compares the variance within clusters to that of a random distribution of mock data. The results suggest an optimal cluster number of either 3 or 4 (Supplementary Figure 13A).

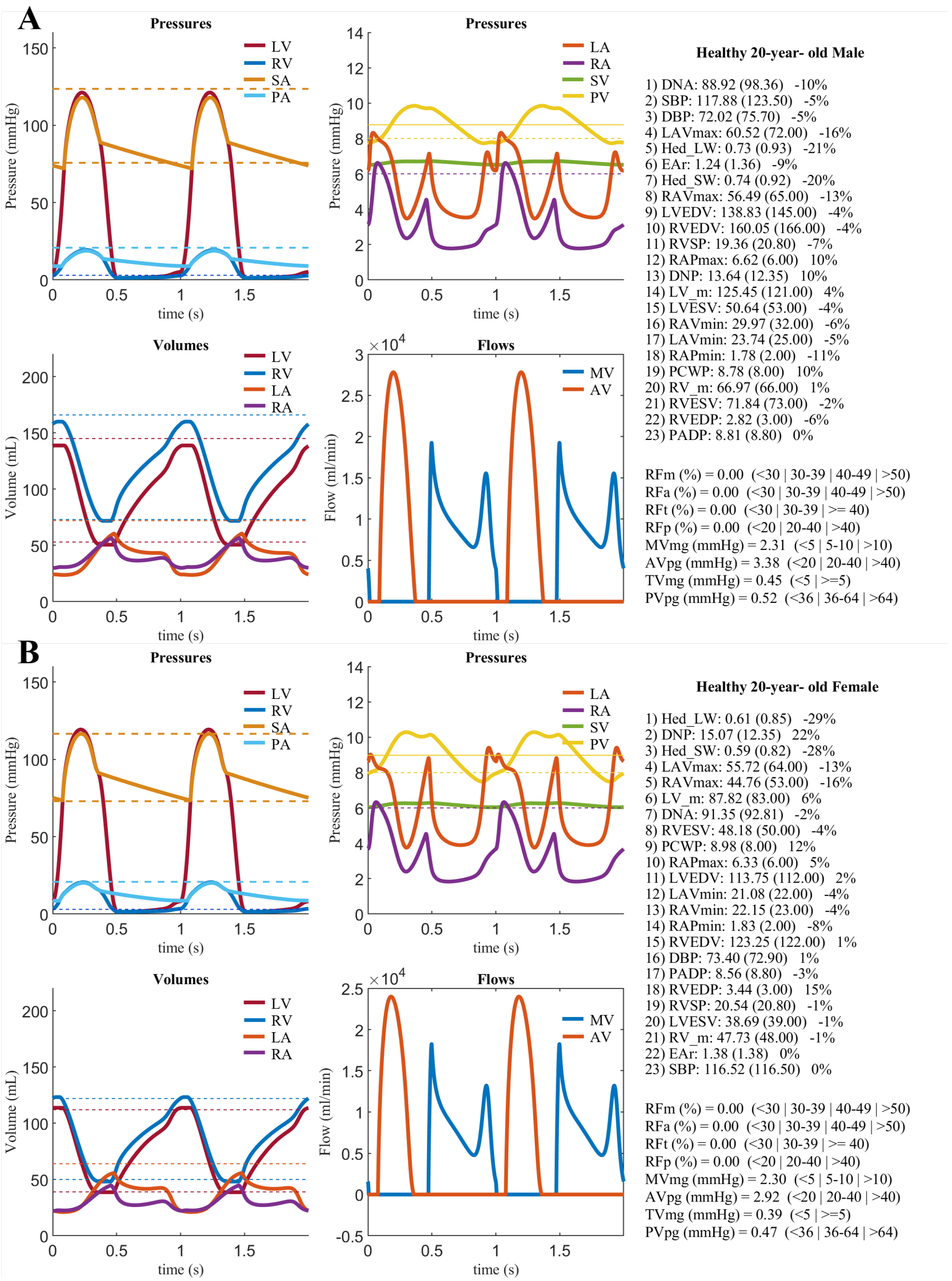


***Supplementary Figure* 12: Simulation outputs for a normal 20-year-old (A) male and (B) female**
Targeted clinical measurements are shown on the right side of each panel. Values in parentheses represent average measurements for healthy 20-year-olds, obtained from previous literature [9–12]. Other abbreviations are defined in Figure 2.

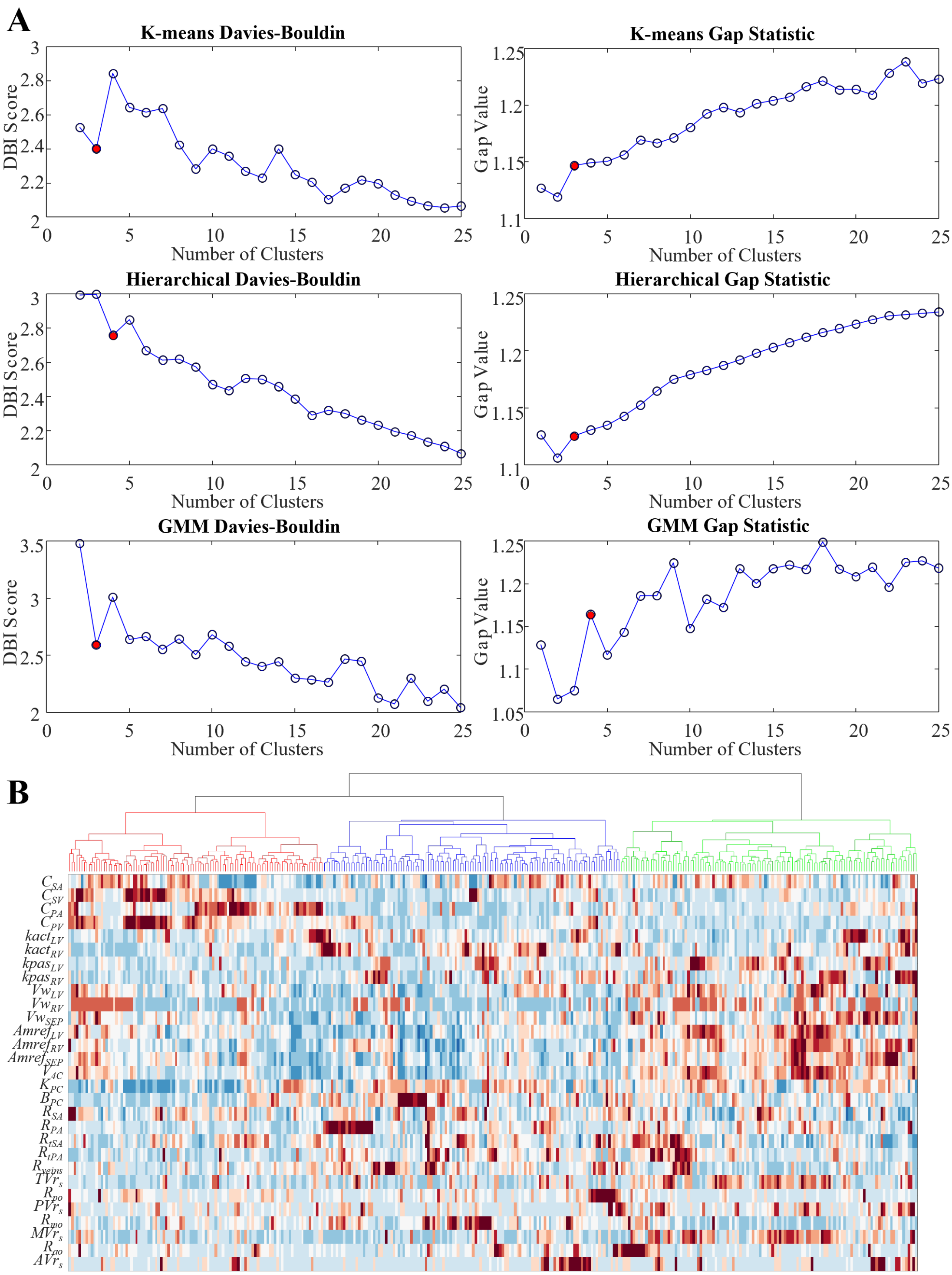


***Supplementary Figure* 13: Optimization of cluster numbers**

(A) Clustering performed using K-means, Hierarchical, and Gaussian Mixture models (GMM), evaluated with the Davies-Bouldin Index (DBI) and Gap statistic. (B) Dendrogram of the hierarchical tree based on functional parameters. Other abbreviations are defined above.

The goal here is to identify distinct and interpretable clusters from a complex, heterogeneous dataset. We aim to isolate a subset of patients who are more distinct from each other. By using multiple clustering methods, individuals consistently grouped into the same cluster across techniques are likely to be more distinguishable. To achieve this, we selected k-means and hierarchical clustering as our clustering techniques, representing one popular linear method and one nonlinear method, respectively.

We did not choose GMM, as it is a soft clustering method that may be more appropriate for mixed data types. We investigated clustering performance with $n = 3$ and $n = 4$ clusters using both k-means and hierarchical clustering. K-means was performed with MATLAB's built-in kmeans function, while hierarchical clustering was conducted using the cluster function, which implements bottom-up clustering. By applying this combined approach, we found that for $n = 3$, 252 patients were consistently assigned to the same clusters across both methods, with 91 inconsistencies. For $n = 4$, the number of inconsistent assignments increased to 106 (Supplementary Figure 14). Based on these results, along with PCA loadings, dendrogram analysis, and ease of interpretability, we selected $n = 3$.

#### 4.2. Strategy of clustering

The subset of 252 patients consistently clustered were used to perform k-means, and the cluster centers were obtained. These patients, along with the cluster centers, were projected into a 2D PCA space (Supplementary Figure 14A). Patients who were not consistently clustered were also mapped onto this space. Using MATLAB's built-in knnsearch function, these patients were assigned to the nearest cluster centers based on Euclidean distance. The final clustering results are visualized in the PCA space (Figure 3A).

The clustering method used may not yield mathematically optimal clusters in terms of within-cluster variance and between-cluster separation. The main challenge is that this heterogeneous disease may not present any clear, distinct clusters. We applied various clustering techniques using baseline clinical characteristics (results not shown), but none produced distinguishable clusters, and they were often difficult to interpret, which contradicts our goals. While we could leverage digital twins to interpret clusters driven solely by clinical characteristics, this approach is indirect, and correlation does not imply causality. In summary, we believe that focusing on a consistent subset leads to more distinct and interpretable clusters, providing mechanistic insights.

#### 4.3. Design of 3D Volcano Plot

To visualize the differences in baseline features across the three groups, we recreated a 3D volcano plot inspired by transcriptomic data [13] in the MATLAB environment. Unlike gene expression data, where features are compared relative to a reference, the baseline features in our study lack such a reference. We redesigned the polar coordinate axes at the base of the cylinder. However, it is actually a Cartesian quadrant, with three vectors representing the three directions. Similarly, each feature has three differences: PG2-PG1, PG3-PG2, and PG1-PG3. These differences are projected onto the corresponding polar axes (Supplementary Figure 4C), and the final position of each feature in this polar coordinate system is determined by summing the vectors along the three axes. Based on the feature's final position, the base of the cylinder is divided into 13 sections. Gray areas indicate differences that are not large enough, while colored regions, as shown in Supplementary Figure 4B, represent varying magnitudes of differences along the three directions.

In practice, we first ensured that our data included both continuous and binary variables. Continuous variables were Z-scored, and binary variables were treated as 0 or 1 (Supplementary Figure 4A). Some features had significant missing data, but we retained all features and performed statistical analysis only on the available data, without imputing missing values. Since the downstream statistics involved variance analysis and chi-squared tests, the number of samples in each group varied dynamically. For continuous variables, Z-scoring was applied, and outlier handling followed the method described in Section 4.1. The calculated differences were projected onto the polar coordinate system shown in Supplementary Figure 4B. For binary variables, the differences were multiplied by 5 before being projected. Features with a projection distance of less than or equal to 1 from the center were considered non-significant and colored gray. The Z-axis represents -log10(P), where P-values for continuous variables come from ANOVA, and for binary variables, from chi-squared tests. P-values were corrected using the Bonferroni method. If $P > 0.05$, the differences were also considered non-significant and colored gray The final results are displayed in a 3D volcano plot (Figure 3D, Supplementary Video 2).

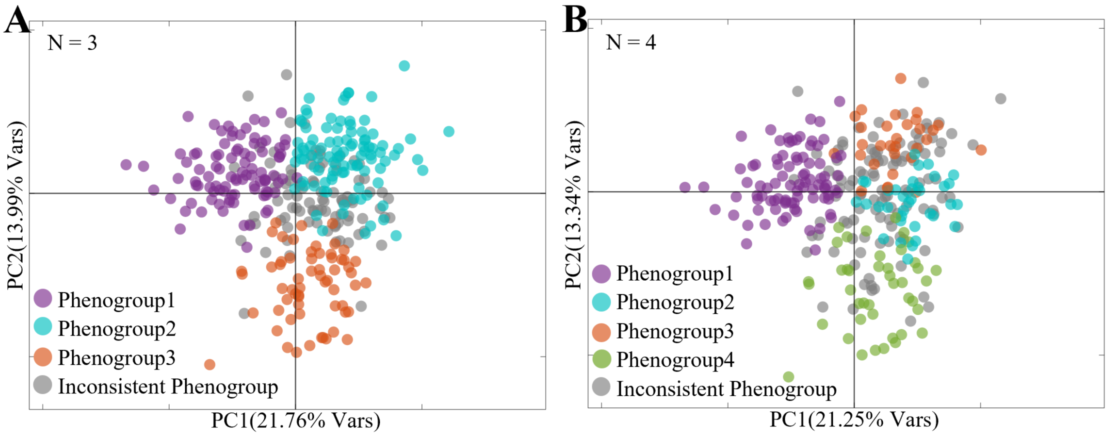


***Supplementary Figure* 14: Optimization of cluster numbers**
Patients were clustered using both K-means and Hierarchical methods. The PCA projection shows consistently clustered patients (colored dots), while inconsistent patients (gray dots) are projected into the same 2D space. (A) Clustering results with N = 3. (B) Clustering results with N = 4.

#### 5. Method S4: Development of the Prognostic Model

Since the UMHS and UW cohorts have different baseline clinical characteristics, we first selected the overlapping features to build the following prognostic models. The prognostic models were developed using R version 4.4.1.

##### 5.1. Random Survival Forests

The Random Survival Forests (RSF) model was trained in R 4.4.1 using the randomForestSRC and VIM packages. Features were divided into three subsets: clinical characteristics, digital twins, and a combination of both. Features with more than 20% missing data were removed (14 out of 146), while the remaining missing data were imputed using the KNN function from the caret package, with $K = 5$. Feature selection was initially based on the variable hunting method [14], followed by stepwise selection of additional features that contributed most to the out-of-bag (OOB) C-index. The selection process was repeated 25 times, and features were ranked by selection frequency and variable importance. Features selected 20 or more times were treated as mandatory, while those selected between 5 and 20 times were flexibly combined with the mandatory features. The final feature set was chosen based on the combination with the highest average OOB C-index. Since a significant number of features are derived from both simulation outputs and clinical characteristics, which are likely representing the same underlying factors (as shown in Supplementary Figure 1), when both types of features are selected, we retain only the one with the higher variable importance.

The hyperparameters, including the number of trees, the number of variables randomly selected at each split, node size, maximum node depth, and split rule, were optimized using a grid search based on the OOB C-index. The sampling type was set to "without replacement", with the sample size determined as 63.2% of the total data. After selecting the optimal hyperparameters and features, the RSF model was trained 25 times to evaluate the uncertainty in the performance metrics. The metrics used include OOB C-index, time-dependent ROC, and integrated AUC (iAUC). These metrics were calculated using the ‘survAUC’ package.

The extended *MAGGIC* score was trained and evaluated using a similar strategy, with the key difference being that variable hunting started with the *MAGGIC* score. Additional features were only selected from the digital twins data.

##### 5.2. Cox Proportional Hazards Model

The Cox proportional hazards model was also built using R 4.4.1 with the glmnet and survival packages. The data preprocessing followed the same approach as described in Section 5.1. Similarly, subsets based on clinical characteristics, digital twins, and their combination were obtained. Feature selection began with univariate Cox regression, selecting features whose hazard ratios (HR) were significantly greater or less than 1. Lasso regression

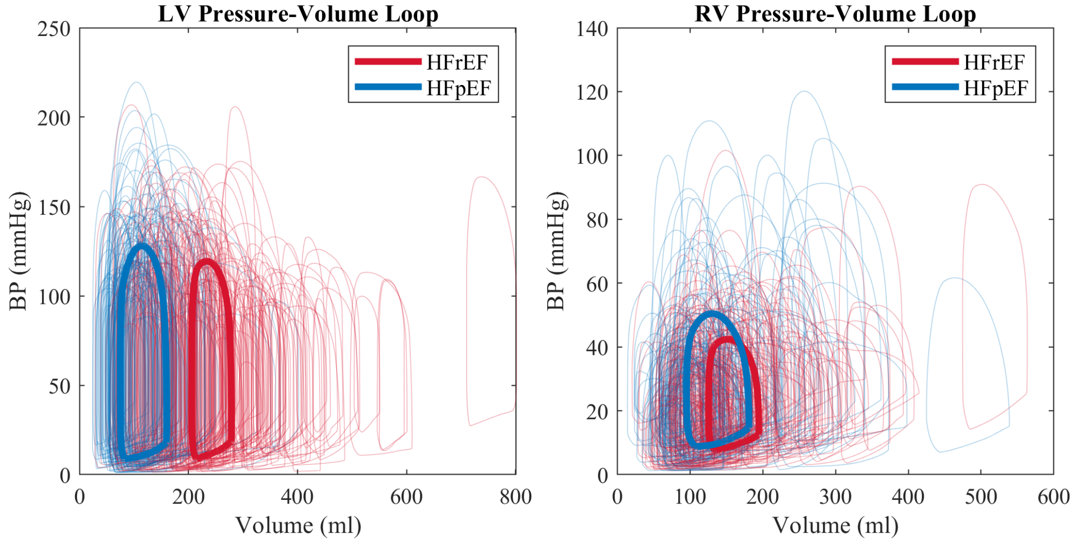


***Supplementary Figure* 15: Pressure-volume loop of the (A) left ventricle (LV) and (B) right ventricle (RV)** Each loop represents a patient from the UMHS cohort. The loops with thicker lines indicate the average for either HFrEF or HFpEF groups.

with 10-fold cross-validation was then applied to further refine the selection and penalize collinear features[15]. The model performance was assessed using the same evaluation metrics as in Section 5.1.

#### 6. Results S1: Performance of Computational Model and Physiological Interpretation of Digital Twins

The relationships between simulated and measured variables for all subjects from UMHS cohort are illustrated in Supplementary Figure 1, revealing that digital twins robustly capture the diverse cardiovascular hemodynamic characteristics present in HF patients. Moreover, in many cases a mismatch between data and model reveals an inconsistency in the data rather than a shortcoming of the model. For example, measured EF's are not necessarily consistent with data on cardiac output, heart rate, stroke volume, end-disatolic volume, and valve regurgitation, while the overall model fit represents an optimal compromise between all of the available data. In addition, the digital twin provides an model-based estimate of key hemodynamic measurements sometimes included in the RHC report. Simulation outputs show correlation with RHC report estimates (Supplementary Figure 2). Similar to EF measurements, model-based outputs are likely to be more accurate than estimates in the RHC report, which do not correct for data inconsistencies and are based on a less comprehensive picture of the patient compared to the digital twin representation.

Digital twins have been further applied to interpret cardiovascular physiology in HF patients. The physiological functional parameters for these patients were standardized and expressed as relative values compared to those of ideal 20-year-old healthy subjects, with female patients compared to an ideal healthy female and male patients compared to an ideal healthy male (Supplementary Figure 12). Consistent with current understanding, both LV and RV contractility are lower in HFrEF than in HFpEF (Supplementary Figure 3A). Notably, HFpEF in this cohort also exhibits reduced LV contractility compared to normal subjects, while a significant portion of HFpEF patients show increased RV contractility (Supplementary Figure 3A).

Examining ventricular power output through simulated pressure-volume loops (Supplementary Figure 15) reveals that both LV and RV power outputs are lower in HFrEF than in HFpEF, reflecting changes in contractility (Supplementary Figure 3A). Specifically, LV power output in HFrEF patients is lower than in normal subjects, while in HFpEF patients, LV power output remains preserved (Supplementary Figure 3A). Additionally, RV power output is significantly increased in both HFrEF and HFpEF (Supplementary Figure 3A).

The passive stiffness of both LV and RV, as quantified through the digital twins approach, is elevated in HFrEF and HFpEF compared to normal subjects, with LV passive stiffness being significantly higher in HFrEF than in HFpEF (Supplementary Figure 3A). The surface areas of the LV, RV, and septum in HFrEF are significantly larger than in HFpEF and normal subjects, indicating a greater degree of dilation in HFrEF hearts (Supplementary Figure 3B). In contrast, the free wall volume does not differ substantially, suggesting that while the heart enlarges, wall thickness decreases, consistent with the pathology of HFrEF (Supplementary Figure 3B). Interestingly, pericardial constraint is markedly elevated in both HFrEF and HFpEF compared to normal subjects, with a greater increase observed in HFpEF, potentially implicating pericardial involvement in HFpEF etiology[16] (Supplementary Figure 3C).

In systemic circulation, the compliance, resistance, and transmural resistance of systemic arteries in both HFrEF and HFpEF are comparable to those in normal subjects (Supplementary Figure 3D). However, HFrEF exhibits slightly but statistically significant increases in both compliance and resistance compared to HFpEF (Supplementary Figure 3D). The compliance of systemic veins in both HFpEF and HFrEF is lower than in normal subjects, with HFrEF showing significantly higher compliance than HFpEF (Supplementary Figure 3D).

In this digital twin analysis, identical resistance values were assumed for pulmonary and systemic veins, resulting in a single value for venous resistance. As a result, venous resistance in HFrEF and HFpEF shows no significant differences and is comparable to normal subjects (Supplementary Figure 3E). In pulmonary circulation, the compliance of both pulmonary arteries and veins is lower than in normal subjects, while the resistance and transmural resistance of pulmonary arteries are elevated in both HFpEF and HFrEF (Supplementary Figure 3E). No significant differences are observed in pulmonary circulation functional parameters between HFpEF and HFrEF (Supplementary Figure 3E).

#### 7. Results S2: Key Feature Differences Across Phenogroups Revealed by 3D Volcano Plot

As mentioned in the Method S3, A modified 3D volcano plot is used to visualize features that show significant differences between phenogroups, as well as how their values differ between phenogroups 1, 2, and 3 (Figure 4D, Supplementary Video 2, Supplementary Figure 4). Each dot in the plot represents a feature, where gray dots indicate insignificant differences, and colored dots indicate significant differences. Each color group has its own meaning showen in Supplementary Figure 4B. Representative features from each color group are labeled and graphically compared between phenogroup populations(Supplementary Figure 16).

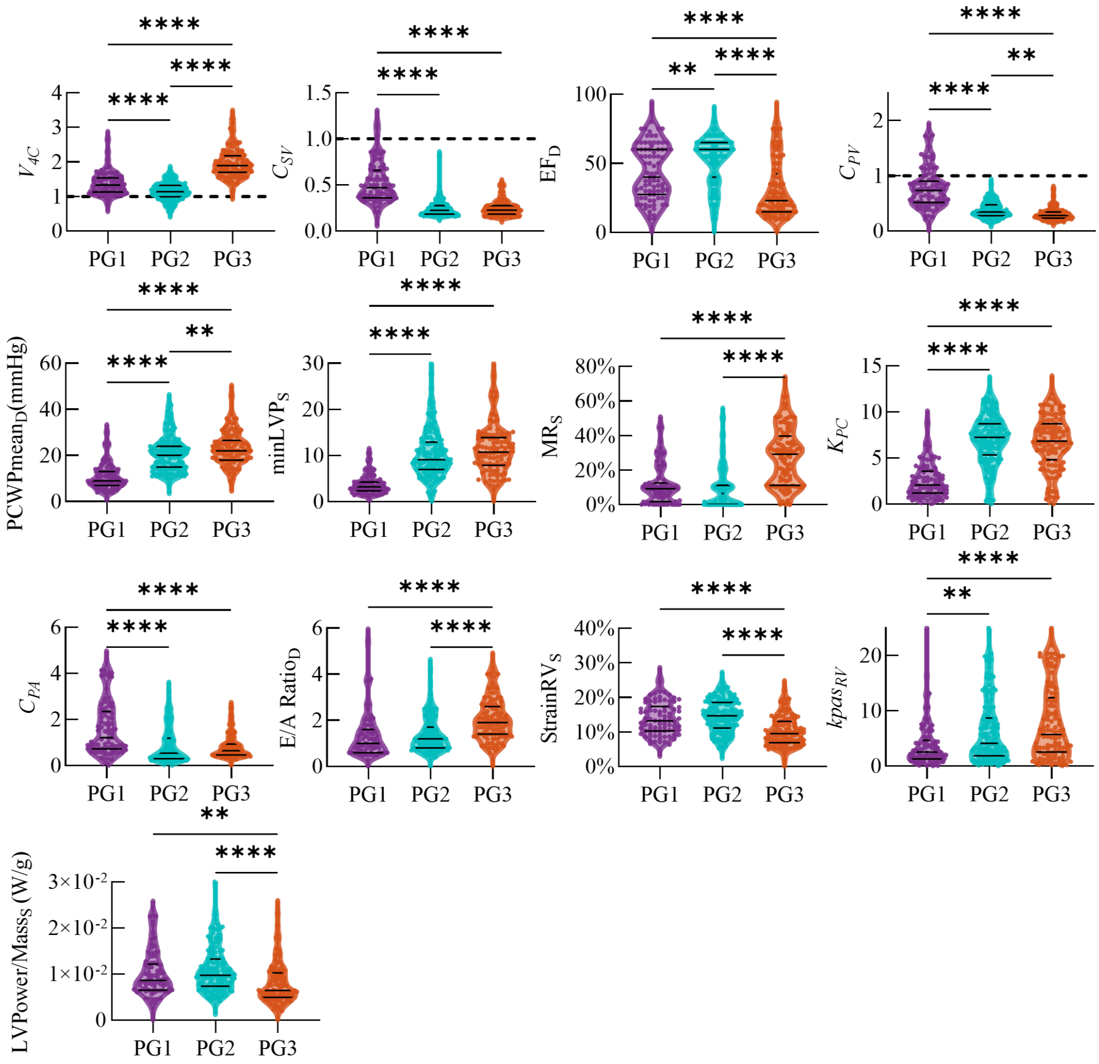

***Supplementary Figure* 16: Feature differences across phenogroups**
All panels show quantitative results corresponding to Figure 3D. This figure highlights representative features from each color group in the 3D volcano plot, aligned with the plot's design as emphasized in Supplementary Figure 4B,C.

In addition to the parameters discussed in the main results, Phenogroup 3 demonstrates the highest RV passive stiffness ($kpas_{RV}$), with Phenogroup 2 also exhibiting higher RV passive stiffness than Phenogroup 1. Phenogroup 1 generally has the lowest RV passive stiffness and the lowest pericardial constraint (Figure 3F). Phenogroup 3 is characterized by the lowest ejection fraction (EF) and body mass index (BMI), the highest prevalence of dilated cardiomyopathy, and elevated levels of B-type natriuretic peptide (BNP). Phenogroup 2 has the highest EF and the greatest incidence of pulmonary hypertension, while Phenogroup 1 presents the lowest BNP levels and an EF comparable to that of Phenogroup 2 (Figure 3E). Hemodynamic and heart morphology differences across phenogroups are described in the main results. Furthermore, Phenogroup 3 patients exhibit the most severe mitral valve regurgitation (MR) and tricuspid valve regurgitation (TR). Phenogroup 2 also shows elevated RV power output and RV free wall stress peak, similar to levels observed in Phenogroup 3 when compared to Phenogroup 1. However, the LV power/mass ratio [17] in Phenogroup 2 remains preserved and is comparable to that of Phenogroup 1 (Figure 4G).

Outcome differences were assessed not only for primary composite endpoints and all-cause mortality but also for LVAD implantation and heart transplantation. Similar associations between phenogroups and outcomes were

**Supplementary Table 4**
**Association between phenogroup membership and the risk of different adverse clinical outcomes using a multivariable adjusted Cox proportional hazard analysis**

| | Phenogroup 1 ($n$ = 83) | | Phenogroup 2 ($n$ = 151) | | | Phenogroup 3 ($n$ = 109) | | |
|---|---|---|---|---|---|---|---|---|
| | Events | Reference | Events | HR (95% CI) | $P$ | Events | HR (95% CI) | $P$ |
| LVAD implantation | 1 (1.2%) | Ref. | 2 (1.3%) | 0.93 (0.08–20.53) | 0.95 | 14 (12.8%) | 11.87 (2.27–218.8) | 0.02 |
| Heart transplantation | 2 (2.4%) | Ref. | 1 (0.7%) | 0.18 (0.01–2.04) | 0.18 | 5 (4.6%) | 2.82 (0.53–22.37) | 0.25 |

CI, confidence interval; HR, hazard ratio.
Outcomes were adjusted for age, sex, smoking status, race, history of diabetes, hypertension, chronic obstructive pulmonary disease (COPD), baseline creatinine levels, usage of angiotensin-converting enzyme inhibitors (ACEi), angiotensin II receptor blockers (ARBs), and beta blockers, as well as for whether the patient had a first diagnosis of heart failure within the past 18 months.

observed with LVAD implantation (Supplementary Figure 5A). However, no significant association was found between phenogroups and heart transplantation, likely due to the limited number of patients reaching this outcome (Supplementary Figure 5B). Prognostic differences across phenogroups, observed in the overall HF cohort, were also apparent in the HFrEF subgroup but became insignificant in the HFpEF subgroup (Supplementary Figure 5C). This may be due to the majority of HFpEF patients clustering into Phenogroup 2, which limits statistical power.

A multivariable Cox proportional hazards model was also applied to assess the association between phenogroups and both LVAD implantation and heart transplantation, using Phenogroup 1 as the reference group. Phenogroup 3 was significantly associated with a higher risk of LVAD implantation (HR 11.87, 95% CI 2.27-218.8) compared to Phenogroup 1, but not with heart transplantation (Supplementary Table 4). No significant associations were observed for Phenogroup 2.

#### 8. Results S3: Prognostic Model

A conventional Cox proportional hazards model was also developed. Feature selection was conducted using Lasso-Cox regression with 10-fold cross-validation, as detailed in Supplementary Table 5. Using the UMHS cohort as the training set, the C-indices for Cox models based on clinical characteristics, digital twins, and combined features were 0.777, 0.669, and 0.789, respectively. The corresponding iAUC values were 0.807, 0.707, and 0.829 (Supplementary Figure 17A). However, in external validation, the performance of all Cox models declined, with C-index values dropping to 0.624, 0.680, and 0.670, respectively, and iAUC values decreasing to 0.606, 0.674, and 0.662 (Supplementary Figure 17B).

The RSF models were also valadated by only 32 HFpEF patients from UM cohort. The RSF model based on combined features still has the highest C-index (0.634), outperforming models using either clinical characteristics (0.556) or digital twins (0.531). But the overall predictive power is low (Supplementary Figure 6A). The time-dependent ROC results are consistent with these conclusions, with iAUCs of 0.549, 0.523, and 0.634, respectively (Supplementary Figure 6B). In terms of prediction within 1 year, the RSF models based on combined features continue to show the highest performance, with AUCs of 0.674 at six months and 0.722 at one year(Supplementary Figures 6C,D).

For the two deceased UW cohort participants analyzed, their risk scores were consistently elevated across all prediction models: the RSF models based on clinical data alone, digital twin data alone, and a combination of both. Among these, the combined model yielded the most pronounced distinction between deceased and surviving subjects, placing the deceased at the top risk rankings—1st and 3rd positions. This separation was less marked with the clinical-only model (rankings of 1st and 4th) and with the digital twins-only model (rankings of 1st and 9th), as illustrated in Supplementary Figure 7.

#### 9. Results S4: Utilizing Longitudinal Data

Most previous phenomapping studies lack longitudinal data, limiting their ability to capture trends in key variables and potentially missing important nuances in phenotype classification. While the current study may also face this limitation, we believe that, unlike many clinical variables that fluctuate rapidly, our digital twin signals are relatively stable over time. Therefore, despite not using longitudinal data, we expect the phenogroups derived from our digital twins to remain consistent and stable. In the UMHS cohort, however, we identified seven patients with two

**Supplementary Table 5**
**Selected variables for Cox proportional hazards model**

| Clinical Characteristics (n = 19) | Digital Twins (n = 10) | Combined (n = 20) |
|---|---|---|
| NYHA Class | **E/A Ratio** | NYHA Class |
| Cardiac Output | **Tricuspid Valve Regurgitation Fraction** | Heart Rate |
| E/A Ratio | *RV Passive Stiffness* | *E/A Ratio* |
| Hemoglobin | *Transmural Resistance of Pulmonary Arteries* | Diabetes Mellitus |
| Diabetes Mellitus | **LV Power Output** | Atrial Fibrillation |
| Age | **PASP** | Hemoglobin |
| Heart Rate | *LV Active Contractility* | Age |
| BMI | *Compliance of Pulmonary Arteries* | Alcohol Use |
| Alcohol Use | **LVEDP** | End Stage Renal Disease |
| Pulmonary Hypertension | **RVEDP** | Drug Use |
| Atrial Fibrillation | | *Transmural Resistance of Pulmonary Arteries* |
| BSA | | Pulmonary Hypertension |
| Drug Use | | BNP |
| BNP | | BMI |
| Tricuspid Valve Regurgitation | | **LV Power Ouput** |
| End Stage Renal Disease | | Tricuspid Valve Regurgitation |
| Usage of Diuretics | | *RV Passive Stiffness* |
| Maximum RA pressure | | Cardiac Output |
| RVSP | | Usage of Diuretics |
| | | *LV Active Contractility* |

Italics represents functional parameters from digital twins, bold represents simulation outputs from digital twins, and underlined represents comorbidities. Normal text without any formatting represents other clinical characteristics. Abbreviations are as defined above.

phenotyping time points and observed their phenogroup transitions. Although only three of the seven patients remained in their original phenogroups, most transitioned from higher to lower risk groups (e.g., from phenogroup 3 to 2 to 1), with the exception of one patient who transitioned from 1 to 2. Notably, all seven patients survived beyond five years, aligning with our interpretation of the phenogroup's association with the risk of the primary composite endpoint (Supplementary Figure 18).

#### 10. Discussion, Limitations and Future Direction

HF is a highly heterogeneous and complex condition, and even at the individual level, patient care generates vast amounts of clinical data. This sheer volume of data makes it challenging to capture the key features of a patient's condition, complicating efforts for individualized management. By implementing a mechanistic computational model to identify digital twins, we provide an integrated and comprehensive assessment of each patient's cardiovascular physiology. This model integrates heart imaging and hemodynamic data into patient-specific functional parameters, simulating cardiovascular system behavior. Although the complexity of the implemented mechanistic computational models introduced a form of bias, this bias is more direction-neutral rather than being biased in any specific direction.

Although we observed that outcomes worsened progressively from Phenogroup 1 to Phenogroup 3—with Phenogroup 3 exhibiting both structural and functional changes, and Phenogroup 2 showing only functional changes compared to Phenogroup 3—this finding does not imply that these phenogroups represent a sequential progression of heart failure. Instead, this observation likely reflects the heterogeneity of heart failure, where patients with distinct key features exhibit different outcomes.

Our cohort selection, which focuses on HF patients undergoing RHC, TTE, and CMR within a limited time window, is likely biased towards patients with more severe and complex conditions. This increased severity and heterogeneity compared to the general HF population has been further validated by our clinical team. Consequently, although the conclusions from the predictive AI component have been externally validated in another prospective cohort, the generalizability of the approach warrants further testing. Additionally, the TriSeg heart model used in our study requires right ventricular geometrical information, yet many patients lack data on the right ventricular free wall mass/thickness. Although we made reasonable assumptions based on patients with complete datasets, this limitation

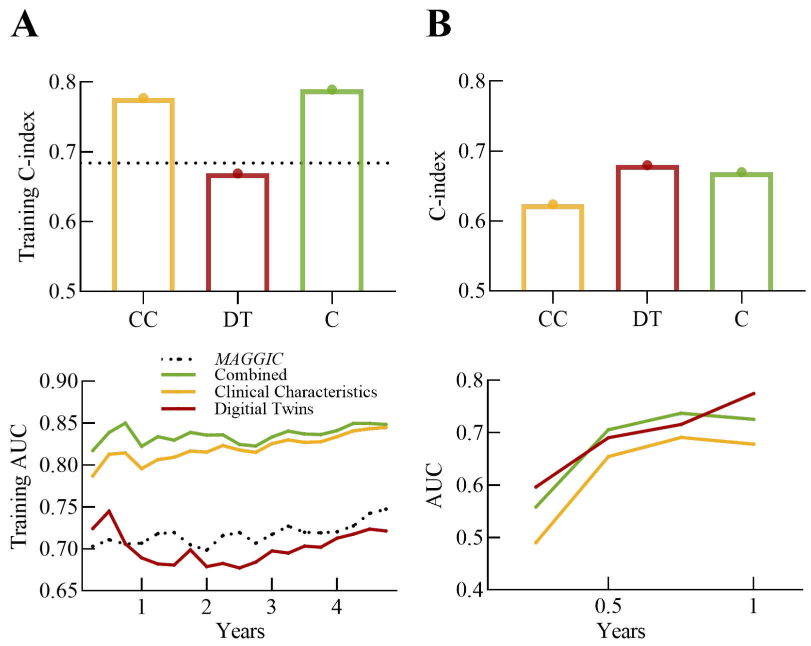


***Supplementary Figure*** **17: Cox proportional hazard regression model performance**
(A) C-index and time-dependent AUC for the training set (UMHS cohort) and (B) external validation set (UW cohort).

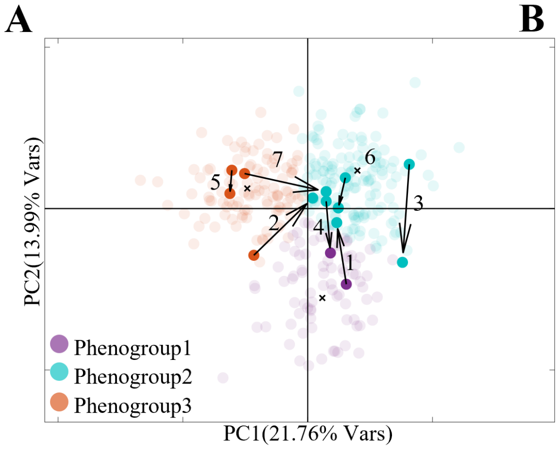


**B**

| Pat No. | Duration between 2 windows | Outcomes |
|---|---|---|
| 1 | 651 days | Alive |
| 2 | 1899 days | Died at 1791 days after second window |
| 3 | 352 days | Alive |
| 4 | 71 days | Alive |
| 5 | 1386 days | Alive |
| 6 | 189 days | Alive |
| 7 | 726 days | Have heart transplantation at 8 days after first window and alive at the end of study |

***Supplementary Figure*** **18: Transition of phenogroups over time**
(A) Phenotype mapping results for 7 patients with two phenotyping windows, showing transitions between phenogroups over time. (B) Corresponding clinical outcomes for the 7 patients.

still weakens the accuracy of the simulations for right ventricular mechanics. Furthermore, the primary composite endpoints in the prognostic AI model differ between the training and external validation datasets. The training dataset uses all-cause mortality, LVAD implantation, and heart transplantation as endpoints, while the external validation set includes all-cause mortality and rehospitalization. This discrepancy arises because using a uniform definition would result in too few events for meaningful statistical analysis in testing set (Supplementary Figure 7). Additionally, the lack of longitudinal data is a significant limitation, as this information is crucial for a comprehensive further analysis.

To advance the digital twin approach, future studies should be conducted in multi-center prospective cohorts to increase the generalizability and applicability of this approach to a broader HF population. Moreover, the model should be refined to accommodate input data lacking right ventricular geometrical measurements, which are often challenging to obtain in clinical practice. Furthermore, integrating longitudinal information into the digital twins would allow for more dynamic and varied clinical scenarios, improving the understanding and management of patient-specific disease progression. In addition, we also envision incorporating exercise hemodynamic measurements to create exercise-state digital twins. This could be particularly beneficial for the early diagnosis and management of HFpEF, providing deeper insights into how patients' cardiovascular systems respond to physical stress. These improvements have the potential to overcome the current study's limitations, paving the way for more robust and comprehensive utilization of the combination of digital twins and AI in heart failure management.
